# One-to-one peer support work in mental health services: systematic review and component network meta-analysis

**DOI:** 10.64898/2026.08.06.26359669

**Authors:** Yasuhiro Kotera, Christopher Newby, Ashleigh Charles, Benjamin-Rose Ingall, Yu Uneno, Fiona Ng, Alex J Sutton, Laura J Gray, Ellesha A Smith, Emma Watson, Larry Davidson, Alan Simpson, Steve Gillard, Bernd Puschner, Sean A Kidd, Candelaria Mahlke, Rebecca Nixdorf, Lisa Brophy, Catherine Brasier, Alison Ashmore, Scott Pomberth, Toshi A Furukawa, Mike Slade

## Abstract

One-to-one peer support is widely used in mental health services, but the components associated with better outcomes remain unclear. We systematically reviewed randomised controlled trials and conducted additive component network meta-analyses to identify which components of one-to-one peer support worker interventions were associated with outcomes for adults using mental health services. CINAHL Ultimate, Embase, MEDLINE, PsycINFO, CENTRAL, ClinicalTrials.gov and ISRCTN were searched, supplemented by citation tracking, previous reviews and expert consultation. Interventions were coded for seven components: Training and development, Maintaining peer support worker wellbeing, Relationship-building, Social support, Emotional support, Practical support and Cultural adaptation. The review followed PRISMA-NMA reporting guidance and was registered with PROSPERO (CRD42022355291). Thirty-six trials randomised 6,645 participants across nine countries. Only quality of life and recovery yielded estimable component effects at one or more follow-up points. For quality of life, Practical support had a positive incremental estimate at 3 months (standardised mean difference 0.52, 95% confidence interval 0.17 to 0.87); no component showed clear evidence of benefit at 6 months; and at 12 months Social support had a positive estimate (1.57, 0.12 to 3.01), whereas Maintaining peer support worker wellbeing had a negative estimate (-1.66,-3.05 to-0.28). These estimates were not consistent across follow-up points. For recovery, Relationship-building had positive estimates at 6 months (0.90, 0.03 to 1.78) and 12 months (0.50, 0.29 to 0.72). Networks were sparse and often disconnected, and additivity could not be tested in disconnected networks. Current trials do not permit definitive prioritisation of peer-support components. Relationship-building was the most consistent candidate component, but all findings remain provisional. Future trials should prospectively specify, manipulate and measure component delivery.

## INTRODUCTION

Mental health peer support (PS) aims to promote mental health recovery through a relationship between mental health service users and PS Workers (PSWs)(1). PSWs are people with lived experience of using mental health services and/or of mental health symptoms, who work in the mental health system to support service users. The relationship is characterised by mutuality and connection (2), which can be achieved by the shared experience of mental health symptoms between a service user and a PSW (3). Sharing personal experience is a defining feature of the PSW role, distinguishing PSWs from other professional groups.

Since the 1990s, PSW roles have been implemented in many countries (4). It is now considered a central component of behavioral and mental health care systems in several countries, including the US, Canada, Australia and New Zealand, while still requiring clearer evidence on fidelity, implementation and mechanisms (5–7). There were more than 10,000 PSWs estimated in the United States in 2006 (8). In England, PS is incorporated in the national mental health implementation plan, which highlights the need for more PSWs (9). Active use of PS in Africa indicates the relationship-based work of PS is also acceptable to people oriented to collectivistic culture (10). A recent multicenter randomized controlled trial (RCT) across high-, middle-and low-income countries also reported benefits of PS for social inclusion, empowerment and hope among people with severe mental health conditions (11). The person-centered value in PS aligns with the current global emphasis on promoting human rights (12). Furthermore, PS is implementable in countries across different socioeconomic levels (13).

Evidence for PS has expanded across effectiveness, implementation and experiential literature (14), including recent umbrella reviews highlighting benefits particularly for recovery-oriented outcomes (15, 16). A meta-analysis on 19 RCTs of one-to-one PS concluded that PS was effective for recovery (SMD=0.23) and empowerment (SMD=0.23) (3). Another meta-analysis on eight group-based PS RCTs identified effectiveness for recovery and psychiatric symptoms (17). Little evidence was reported on hospitalization, overall symptoms, and satisfaction with services in a meta-analysis focused on people with severe mental illness, however some evidence on hope, recovery and empowerment was found (18). A systematic review on digital PS, where PS was implemented live or automated using technology, identified that digital PS is feasible, acceptable and effective (19).

A qualitative meta-synthesis reported that service users regard their PSW as a role-model, which strengthens service users’ hope and motivation for recovery (20). Another meta-synthesis found that PS changes service users’ identity positively, supported by mutual relationships and story sharing by the PSW (21). A longitudinal study identified that a PSW’s story sharing helps service users transform fears into hope (22).

Moreover, PS is effective for diverse populations. PS has been assessed positively with older adults, youth, LGBTQ+ people, persons with disabilities, and those in a forensic context (23–25). It is also increasingly discussed for people with co-occurring mental health and substance use disorders, where peer specialists may contribute to social inclusion through lived experience, authenticity and social engagement (26). A wide range of population groups can benefit from PS (27).

Given this strong evidence base for PS, it is important to understand its mechanisms of action. It is clearly a complex intervention, with some arguing that this complexity means PS work cannot be evaluated as a single intervention (28). However, mixed-methods approaches to develop a theoretically-informed PS intervention have been published, e.g. the development of the ENRICH Intervention using systematic reviews, expert panels, consensus methods and co-production (29). Another example is the development of the training programme for the UPSIDES Intervention through systematised review and international Delphi consultation (30). More broadly, realist evaluation has identified five contextual factors that together form the APPEAR framework (Accepting, Personalised Practice, Empowering, Available and Reciprocal) (31). We synthesised current evidence to develop a PS component typology identifying seven components (32). This typology allows investigation of component-level impact of PSW on outcome.

The aim of this study was to identify which components, and combinations of components, in one-to-one PS are associated with outcomes for adults using mental health services.

## METHODS

### Design

A systematic review including component network meta-analysis (CNMA) was performed. We followed the PRISMA Extension Statement for Reporting of Systematic Reviews Incorporating Network Meta-analyses of Health Care Interventions (S1 Supporting Information)(33). The protocol was pre-registered at PROSPERO (CRD42022355291).

### Data search

Twenty-six studies were identified in our previous systematised review to identify PS components (32) (S2 Supporting Information). The sources for the 26 studies were (a) a recent systematic review of one-to-one PS for adults (3), (b) database searches on Medline and PsycINFO (the last search on 23rd August 2022), and (c) expert consultation (21 researchers and practitioners specialised in PS; January 2023). Two reviewers, YK and AC, independently assessed the eligibility of 10% of the studies to establish concordance. The rest were assessed by YK. In case the two reviewers did not reach a consensus, a third reviewer was nominated for consultation (MS), however the consultation was not needed.

To update our searches, three data sources were used: databases, clinical trial registers and an umbrella review (15). First, Medline and PsycINFO were searched for publications from 2022 to 1st June 2023. Additionally, EMBASE and CINAHL Ultimate were searched for publications published until 1st June 2023. The same search strings were used as our previous systematised review (32) (S3 Supporting Information). Second, three clinical trial registries (CENTRAL, ClinicalTrials.gov and ISRCTN) were searched, which were supplemented with Google Scholar (the first 100 results for *‘“mental health”, “peer support”, AND “RCT”’* in the incognito mode) (S4 Supporting Information). The last search was conducted on 11th June 2023. YK searched both databases and trial registers, with consultation by the subject librarian AA. Third, an umbrella review (15) was published during our study, and was reviewed for includable studies. YK and AC independently assessed the eligibility of additional studies.

### Study selection

The eligibility criteria are presented in Table 1.

**Table 1.** Eligibility criteria for the included studies.

|  | **Inclusion criteria** | **Exclusion criteria** |
| --- | --- | --- |
| **Population** | Adults (16 years or over) using mental health services | Children and adolescents who are 15 years or younger<br>Adults (16 years or over) not using mental health services |
| **Intervention** | One-to-one mental health peer support delivered in person or online/remotely | Group-based peer support<br>One-to-one peer support targeting non-mental health outcomes, such as drug/alcohol or breast-feeding |
| **Comparator** | All types |  |
| **Outcomes** | Main: Health-related quality of life, which includes outcomes such as quality of life, wellbeing, psychiatric symptoms, hope, and social support, social functioning<br>Additional: Outcomes that were evaluated in more than 10 included studies from below*:<br>Hospitalization<br>Satisfaction with services<br>Working alliance (clinician rated or patient rated)<br>Engagement with services | Other outcomes |
| **Study design** | Randomized controlled trials, either cluster or individual | Other designs |
| **Other** | Published in a peer-reviewed journal in English language |  |

\* These were outcomes evaluated in a recently published systematic review(3) that shares the same target population as our study: PS for adults using mental health services.

### Data extraction

A data abstraction table was used to extract the following from each study: first author, publication year, country, population, sample size (per arm), descriptions of intervention and control, outcomes and assessments, longest follow-up, type of PS, and organizational support. Two independent reviewers, YK and AC, established concordance in 10% of the included studies. The rest were reviewed by YK. A third reviewer, MS, was nominated for consultation in case the two reviewers did not reach concordance.

### Component identification

Our published component typology of PS identified 16 components (32), which were thematically categorised into seven components by the research team to effectively enable a component network meta-analysis, shown as B to H in Table 2. Each study’s inclusion of components was independently assessed by YK, AC, and BRI. A fourth assessor, MS, was nominated for consultation in case the three assessors did not reach concordance.

**Table 2.** Comparator and PS components, definitions and assessment guidance.

| Label | Component | Definition | Assessment guidance |
| --- | --- | --- | --- |
| A | Comparator arm | The comparator arm against which one-to-one peer support (PS) was evaluated. Comparator arms included care as usual, waitlist, usual services, or an alternative non-peer intervention, depending on the trial. | Coded as “A” for all comparator arms. Comparator arms were not coded as containing peer support components unless they included an active peer support element. |
| B | Training and development | PS workers (PSWs) are trained to offer PS work before, and sometimes during the PS work, by different workers including other PSWs, psychologists, psychiatrists, and research assistants. Training contents include knowledge of mental health, PSW role, ethics and PSW wellbeing. | “Yes” if PSW training or other developmental activities were reported.<br>“No” if none of above were reported |
| C | Maintaining PSW wellbeing | To promote and maintain PSW's wellbeing by PSWs themselves, other PSWs, and their organisations. For example, identifying people PSWs would need to talk to, to ensure that they receive the right support. This component includes PSW meetings and organisational support for PSWs. This component includes PSW supervision. | “Yes” if activities to promote or maintain PSW wellbeing were reported, either (a) by PSWs themselves or (b) by others, such as other PSWs or their organisations.<br>“No” if neither self-directed nor externally provided PSW wellbeing support was reported. |
| D | Relationship-building | Using shared experience, a PSW builds a relationship based on mutuality and connection with a service user. This includes goal-setting (PSWs work with service users to identify and meet service user personal goals and/or shared goals for the PS relationship) and sharing lived experience (PSWs explore methods and strategies for using their own lived experience with service users, including the safe, purposeful, and appropriate use of their own story to benefit others). | “Yes” if relationship-building was specifically described.<br>“No” if relationship-building was not specifically described, i.e. relationship-building was only noted, or not noted at all. |
| E | Social support | To help service users to participate in social activities and/or socialise with others, including going out for coffee/meals, game nights at local recreational centres, hikes or movies. | “Yes” if an intervention had a specific section detailing adaptation for the specific component (e.g. social support).<br>“No” if an intervention did not have a specific section or gave only a brief mention detailing adaptation for the specific component. |
| F | Emotional support | To help service users emotionally feel well, including helping them stay hopeful and motivated, supporting their self-esteem, giving encouragement, or just being present for them. |  |
| G | Practical support | Tangible assistance including support in housing, disability benefits, obtaining medications, groceries, and other daily living activities of daily living. |  |
| H | Cultural adaptation | Modifying PS work to be compatible with the cultural, linguistic, community, service-system, or contextual needs and values of service users. For example, a model used in a different culture has been modified to fit better with the current culture, or a decision of choosing an intervention considered a cultural fit. |  |

Table 2 presents the comparator (A) and seven peer support components (B-H), definitions, and assessment guidance. Components are labelled using letters rather than numbers to avoid confusion with numerical effect estimates in the Results.

### Quality assessment

The Cochrane Risk-of-Bias Tool for Randomized Trials (RoB) was used to assess the risk of bias in the newly included studies (n=17), by two independent reviewers, YK and YU. The rating for the 19 studies included in the previous meta-analysis were taken from the review paper (3). A third reviewer, MS, was nominated to resolve possible disagreement.

## Data synthesis and analysis

### Data preparation

Descriptive statistics were generated for study interventions and comparators, including means and standard deviations (SD) for all outcomes. When alternative summary measures were reported, these were converted to means and SD using Cochrane-recommended formulas (34).

### Outcome domains

The primary outcome was quality of life, because it was a health-related outcome with the largest amount of available data and one of the few outcomes for which component network meta-analysis was feasible. Secondary outcome domains were anxiety, depression, hope, loneliness, recovery, mental health symptoms, hospital use, social outcomes, and work-related outcomes. For each domain, outcome measures at comparable follow-up points were combined into a standardized mean difference (SMD). Four follow-up periods were defined: approximately 3 months (±1), 6 months (±2), 12 months (±3), and 18 months (±3). Data preparation and coding were conducted in Excel and R. Only domains with at least three comparable SMDs at a given time point were included in the quantitative synthesis. Component effects were estimated only when the evidence network allowed components to be separated; otherwise, components were reported as not estimable.

### Network construction

Network geometry was visualized for each outcome domain to determine whether the available evidence formed a closed or open network. Network graphs illustrated the number of studies contributing to each comparison and informed the subsequent analytical model.

### Statistical models

All analyses were conducted using the netmeta package in R. Random-effects models were used to account for heterogeneity across trials, following the CNMA framework proposed by Welton, Caldwell (35) and extended by Freeman, Scott (36). Two analytical stages were conducted: (a) standard network meta-analysis (NMA), which estimated relative effects among the intervention and comparator packages connected within each outcome network; and (b) additive CNMA, which decomposed the packages into their constituent components and estimated the incremental effect of each component, using care as usual as the reference. Effects of component combinations were derived under the additive model. Components that could not be estimated separately because they consistently co-occurred across interventions were amalgamated into a single component.

### Estimation and heterogeneity

Effect estimates are reported as standardised mean differences (SMDs) with 95% confidence intervals (CIs) and associated *p*-values. SMDs of ≥0.3 and ≤0.7 were interpreted as medium effects, and SMDs >0.7 as large effects. Statistical significance was defined as *p*<0.05. Between-study heterogeneity was assessed using τ^2^, I^2^ and Q statistics. Standard NMA was conducted only where the relevant intervention network was connected. Component effects were estimated only where the network structure and distribution of components allowed them to be identified; no fixed minimum number of direct comparisons was applied. Where component effects or heterogeneity statistics could not be estimated because of insufficient network information, they were reported as not estimable.

### Sensitivity and validation

Publication bias was planned to be assessed using funnel plots where sufficient data were available. However, funnel plots were not produced because each network included only one study per comparison, providing insufficient data to assess small-study effects.

## RESULTS

Thirty-six trials were included. Nineteen had been included in the earlier meta-analysis (3); seven additional trials had been identified for the component-typology review (32); and ten further trials were identified for the present review. Of these ten, one was identified by the updated database and registry searches and nine through the 2024 umbrella review (15). Screening the second umbrella review16 identified no additional eligible trial. Figure 1 presents the study flow.

**Figure 1.**
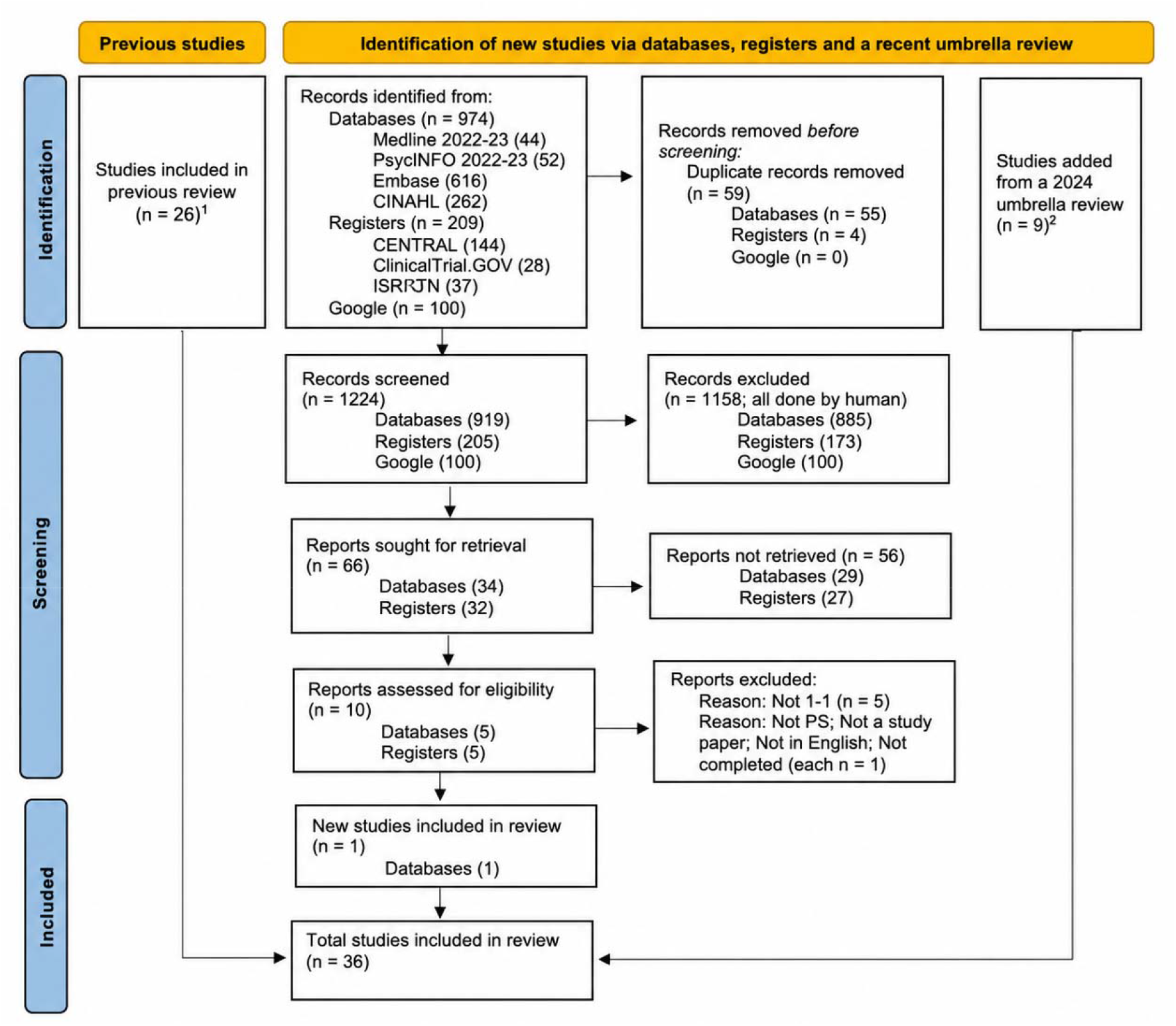
PRISMA flow diagram. Across the 36 studies, 6,645 participants were recruited. The proportions of female, male, and transgender participants were 61.01% (n=4,054), 38.89% (n=2,584), and 0.11% (n=7). Twenty studies (56%) were published between May 2013 and June 2023. All studies were single-country trials. Studies were conducted in the USA (n=20), England (n=5), Canada (n=5), Australia (n=1), France (n=1), Germany (n=1), Japan (n=1), the Netherlands (n=1), and Singapore (n=1). By region, 69% (n=25) were conducted in North America, 22% (n=8) in Europe, 6% (n=2) in Asia, and 3% (n=1) in Oceania. Care as usual (CAU) was the most common comparator (78%, n=28). The mean sample size for the PS groups was 83, with range from 10(37) to 349(38). Most commonly assessed outcomes were quality of life (n=16), psychiatric symptoms including depression and anxiety (n=16), and hospitalization (n=10). Table 3 presents data for all studies (full details in S5 Supporting Information).

**Table 3.** Data abstraction table for included studies (n=36)

| No | Study | Country | Population | Intervention | Control | Outcomes | Longest Follow Up |
| --- | --- | --- | --- | --- | --- | --- | --- |
| #01 | Gillard et al. 2022 (39) | England | Adult psychiatric inpatients with recent prior admission | 294 | 296 (available data 306:267:3) | 1) Subjective quality of life<br>2) Social inclusion<br>3) Hope<br>4) Psychiatric symptom | 12 months |
| #02 | Rohrbach et al. 2022 (40) | Netherlands | Adults with eating disorders | 90 (89:1) Featback (internet intervention) plus weekly PS<br>87 (84:3) Weekly PS only | 90 (88:2) Waitlist<br>88 (82:6) Featback (internet intervention) only | 1) Eating disorder symptomatology<br>2) Anxiety and depression<br>3) General self-efficacy<br>4) Experienced social support<br>5) Motivation, satisfaction, and help-seeking<br>6) Similarity of oneself to PSW | 12 months |
| #03 | Tinland et al., 2022 (41) | France | Adults with schizophrenia, bipolar I disorder, or schizoaffective disorder | 196 (69:127) | 198 (86:112)<br>Total 155: 239 | 1) Therapeutic alliance<br>2) Quality of life<br>3) Mental health symptoms<br>4) Empowerment<br>5) Recovery outcomes | 6 months |
| #04 | Kidd et al. 2021 (42) | Canada | Adults with schizophrenia spectrum illness post-hospitalisation | 41 (19:22) | 22 (6:16) Brief intervention.<br>44 (16:28) CAU | 1) Community ability<br>2) Community engagement<br>3) Symptomatology<br>4) Substance use<br>5) Personal recovery<br>6) Social support<br>7) Quality of life | 6 months |
| #05 | Maru et al. 2021(43) | USA | Adults with psychiatric disabilities using community outpatient services | 83 (43:40) | 83 (42:41) | 1) Vocational and prevocational activity<br>2) Quality of life<br>3) Hope<br>4) Work readiness<br>5) Working alliance | 12 months |
| #06 | Byrne et al., | USA | Adults hospitalised with substance use disorder | 51 (19:32) | 47 (21:26) | 1) Substance use frequency | 6 months |
|  | 2020 (44) |  | complications |  |  | 2) Physical and mental health |  |
| #07 | Ranzenhofer et al., 2020 (45) | USA | Outpatients with anorexia nervosa, bulimia nervosa, or binge-eating disorder. | 20 (20:0) | 18 (17:1)<br>22 (22:0) | 1) Eating disorder symptoms<br>2) Eating disorder quality of life<br>3) Anxiety symptoms<br>4) Depression symptoms | 6 months |
| #08 | Cardi et al., 2019 (46) | England | Adult with anorexia nervosa | 99 (96:3) | 88 (84:4) | 1) Eating disorder symptoms<br>2) Mood<br>3) Quality of life<br>4) Motivation for treatment<br>5) Alliance with the therapist and cognitive and behavioural flexibility | 12 months |
| #09 | Pfeiffer et al., 2019 (47) | USA | Adult psychiatric inpatients at high suicide risk | 34 | 36<br>Total 37:29:4 | 1) Suicide attempts<br>2) Suicidal ideation<br>3) Hopelessness<br>4) Hope<br>5) Belongingness<br>6) Use of services | 6 months |
| #10 | Shorey et al., 2019 (48) | Singapore | Mothers at risk of postnatal depression | 69 (69:0) | 69 (69:0) | 1) Postnatal depression<br>2) Postnatal anxiety<br>3) Loneliness<br>4) Perceived social support | 3 months |
| #11 | Corrigan et al., 2018 (49) | USA | Adult Latinos with serious mental illness | 55 (28:26:1) | 55 (36:18:1) | 1) Recovery<br>2) Personal empowerment<br>3) Quality of life | 12 months |
| #12 | Johnson et al., 2018 (50) | England | Adults receiving crisis resolution team support | 220 (132:88) | 219 (131:87) | 1) Overall psychiatric symptoms<br>2) Social network support<br>3) Recovery<br>4) Satisfaction with services<br>5) Hospitalisation | 18 months |
| #13 | Mahlke et al., 2017 (51) | Germany | Adults with schizophrenia-related, affective, or personality disorders using psychiatric services | 114 (65:49) | 112 (59:53) | 1) Overall psychiatric symptoms<br>2) Quality of life<br>3) Social functioning | 12 months |
|  |  |  |  |  |  | 4) Empowerment<br>5) Hospitalisation |  |
| #14 | Seeley et al., 2017 (52) | USA | Older adults with mild-to-moderate depression and/or anxiety | 31 | 31<br>Total 50:12) | 1) Depression<br>2) Anxiety<br>3) Working alliance | 2.5 months |
| #15 | Yamaguchi et al., 2017 (53) | Japan | Adult psychiatric outpatients receiving case management | 26 (10:16) | 27(12:15) | 1) Overall psychiatric symptoms<br>2) Social Functioning<br>3) Empowerment<br>4) Working alliance | 12 months |
| #16 | Rogers et al., 2016 (54) | USA | Adults court-ordered to treatment following a mental health crisis | 63 (34:29) | 50 (29:21) | 1) Social network support<br>2) Overall psychiatric symptoms<br>3) Recovery<br>4) Quality of Life | 6 months |
| #17 | Salzer et al., 2016 (55) | USA | Adults with schizophrenia spectrum disorder, bipolar disorder, or major depression using community outpatient services | 50 (23:26:1) | 49 (26:23:0) | 1) Quality of life<br>2) Recovery<br>3) Empowerment<br>4) Working alliance | 12 months |
| #18 | Valenstein et al., 2016 (56) | USA | Veterans with depression and prior treatment exposure | 144 (27:117) | 243 (47:196) | 1) Depression<br>2) Mental health and physical health<br>3) Quality of life<br>4) Suicidal thoughts<br>5) Mental health recovery | 12 months |
| #19 | Wroblewski et al., 2015 (57) | Canada | Adults using community mental health services for persistent mental illness, with or without substance use | 12 (11:1) | 9 (7:2) | 1) Quality of life | 6 months |
| #20 | Simpson et al., 2014 (58) | England | Adult acute psychiatric inpatients approaching discharge | 23 (7:16) | 23 (3:20) | 1) Hope<br>2) Quality of life<br>3) Hospitalisation | 3 months |
| #21 | Chinman et al., 2013 (59) | USA | Adult VA intensive case management patients with recent high psychiatric hospital use | 122 (12:110) | 116 (16:100) | 1) Quality of life<br>2) Recovery<br>3) Empowerment<br>4) Overall psychiatric symptoms | QoL - 6 months<br>Other - 12 months |
| #22 | Gjerdingen et al., 2013 (60) | USA | Mothers with depressive symptoms and infants aged 0–6 months | 13 (13:0) | 14 (14:0)<br>Duola group<br>12 (12:0) | 1) Depression and anxiety<br>2) Depression<br>3) Overall health status<br>4) Available support<br>5) Importance of support<br>6) Satisfaction with support<br>7) Illness days | 6 months postenrollment |
| #23 | Proudfoot et al., 2012 (61) | Australia | Adults currently treated for bipolar disorder | 134 (98:36) | 139 (93:46)<br>Psycho-education only.<br>134 (93:41)<br>Brief online texts about bipolar disorder were sent. | 1) Depression and anxiety<br>2) Social functioning<br>3) Empowerment | 6 months |
| #24 | Letourneau et al., 2011 (62) | Canada | Mothers with postpartum depression and infants under 9 months | 27 (27:0) | 33 (33:0) | 1) Maternal-infant interactions<br>2) Depression<br>3) Social support<br>4) Infant development<br>5) Salivary cortisol | 12 weeks |
| #25 | Simon et al., 2011 (63) | USA | Adults currently treated for bipolar disorder | 64 | 54<br>Total 85:33 | 1) Engagement with services | 3 weeks |
| #26 | Sledge et al., 2011 (64) | USA | Adult psychiatric inpatients with repeated recent hospitalisation and severe mental illness | 39 (22:17) | 37 (16:21) | 1) Hospitalisation<br>2) Overall psychiatric symptoms<br>3) Social Functioning<br>4) Hope<br>5) Satisfaction with services<br>6) Social network support<br>7) Wellbeing | 9 months |
| #27 | Dennis et al., 2009 (65) | Canada | Women at high risk of postnatal depression in the first two weeks postpartum | 349 (349:0) | 352 (352:0) | 1) Postnatal depression<br>2) Anxiety<br>3) Loneliness | 24 weeks |
|  |  |  |  |  |  | 4) Health service utilisation |  |
| #28 | Rivera et al., 2007 (66) | USA | Adults with psychotic or mood disorder and repeated recent hospitalization | 70 (35:35) | 66 (31:35) CAU<br>67 (34:33) Clinic-based care by psychologist and social worker | 1) Overall psychiatric symptoms<br>2) Quality of Life<br>3) Social network support<br>4) Wellbeing<br>5) Hospitalisation | 12 months |
| #29 | Sells et al., 2006 (67) | USA | Adults with serious mental illness and treatment disengagement | 68 | 69 (in total 53:84) | 1) Working alliance - client<br>2) Engagement with services | 12 months |
| #30 | Craig et al., 2004 (68) | England | Adults with serious mental illness, poor engagement, and assertive outreach team involvement | 24 | 21 (in total 15:30) | 1) Social functioning<br>2) Social network support<br>3) Hospitalisation<br>4) Satisfaction with services<br>5) Service engagement | 12 months |
| #31 | Davidson et al., 2004 (69) | USA | Psychiatrically stable adults with serious mental illness | 95 PS with \$28 stipend | 70 in passive control with \$28 stipend<br>95 in Partnered by nonconsumer with \$28 stipend. Total gender split 260 (148:112) | 1) Psychiatric symptoms<br>2) Functioning impairment<br>3) Self-esteem<br>4) Social functioning<br>5) Depression<br>6) Wellbeing<br>7) Nonpsychotic Psychiatric Symptoms<br>8) Satisfaction with service | 9 months |
| #32 | Dennis et al., 2003 (38) | Canada | Mothers with postpartum depression 8–12 weeks after singleton birth | 20 (20:0) | 22 (22:0) | 1) Depression<br>2) Self-esteem<br>3) Child-care stress<br>4) Loneliness<br>5) Perceptions of peer support | 8 weeks |
| #33 | Clarke et al., | USA | Adults with severe mental disorder and persistent | 57 (21:36) | 57 (22:35) | 1) Hospitalisation | 6 months |
|  | 2000 (70) |  | psychotic symptoms |  | Non-peer<br>assertive<br>community<br>treatment<br>49 (21:28)<br>CAU |  |  |
| #34 | Hunkeler et al.,<br>2000 (71) | USA | Adult primary care patients with major depression<br>or dysthymia prescribed SSRIs | 62 Telehealth<br>care and PS | 117<br>Telehealth<br>care<br>123 (in total<br>208:94) CAU | 1) Depression and anxiety<br>2) Social functioning<br>3) Satisfaction with services | 6 months |
| #35 | Klein et al.,<br>1998 (37) | USA | Adults with dual diagnosis receiving intensive case<br>management | 10 | 51 (in total<br>16:45) | 1) Hospitalization<br>2) Social functioning<br>3) Quality of Life<br>4) Social network support<br>5) Wellbeing | 6 months |
| #36 | Solomon et al.,<br>1995 (72) | USA | Adults at risk of hospitalization receiving intensive<br>case management | 46 (27:19) | 45 (16:29) | 1) Overall psychiatric<br>symptoms<br>2) Social network support<br>3) Quality of life<br>4) Hospitalisation<br>5) Working Alliance | 24 months |

PS=Peer Support

Eighty-two arms were identified across the 36 included studies, comprising 40 PSW intervention arms and 42 comparator arms. The most frequently identified component was Training and development (B), which was present in 40 arms, followed by Maintaining PSW wellbeing (C) in 33 arms, Social support (E) and Emotional support (F) in 32 arms each, Relationship-building (D) in 28 arms, Practical support (G) in 27 arms, and Cultural adaptation (H) in 15 arms. Full arm-level component coding is presented in S6 Supporting Information.

### Risk of bias

Table 4 presents the risk of bias rating for all 36 studies.

**Table 4.** Risk of bias for 36 included studies.

| No | Random sequence generation<br>(selection bias) | Allocation concealment<br>(selection bias) | Blinding of outcome assessment<br>(detection bias) | Incomplete outcome data<br>(attrition bias) | Selective reporting<br>(reporting bias) |
| --- | --- | --- | --- | --- | --- |
| #01 | L | L | L | L | L |
| #02 | L | L | L | L | L |
| #03 | L | L | L | L | L |
| #04 | L | L | L | U | L |
| #05 | L | L | U | L | L |
| #06 | L | L | L | U | L |
| #07 | L | L | L | L | L |
| #08 | L | L | L | L | L |
| #09 | L | L | L | U | U |
| #10 | L | L | L | L | L |
| #11 | U | U | U | L | H |
| #12 | L | L | L | L | L |
| #13 | L | L | U | L | U |
| #14 | L | U | U | L | L |
| #15 | L | L | U | U | U |
| #16 | L | L | L | U | U |
| #17 | L | L | H | U | U |
| #18 | L | L | U | L | L |
| #19 | U | U | U | H | U |
| #20 | L | U | U | L | L |
| #21 | L | H | H | H | U |
| #22 | L | H | L | L | U |
| #23 | L | L | L | L | L |
| #24 | L | L | L | L | U |
| #25 | L | L | L | L | U |
| #26 | L | U | L | U | H |
| #27 | L | L | L | L | L |
| #28 | U | U | L | L | U |
| #29 | U | U | L | H | H |
| #30 | H | U | H | L | U |
| #31 | U | H | U | U | U |
| #32 | L | L | L | L | U |
| #33 | U | U | L | U | U |
| #34 | U | U | U | L | H |
| #35 | U | U | U | U | L |
| #36 | U | U | U | L | H |

L = low risk of bias; U = unclear risk of bias; H = high risk of bias

Nine trials were assessed as low risk of bias in all five domains. Across the domains, random sequence generation was most frequently assessed as low risk (n=26), followed by incomplete outcome data (n=23), allocation concealment and blinding of outcome assessment (each n=21), and selective reporting (n=16).

### Component network meta-analysis

Hospital use was not meta-analysed because definitions and methods of capture differed substantially across studies. Other planned domains, including anxiety, depression, hope, loneliness, mental health symptoms, social outcomes and work-related outcomes, were not analysed because there were insufficient comparable data at the relevant follow-up points. Quality of life and recovery were retained as the main outcomes for analysis, because these were the only domains with sufficient data for network meta-analysis and, at selected time points, component network meta-analysis.

#### Quality of life: Three-month follow-up

For quality of life at three months, six studies totalling 1,520 participants contributed data to the network (39, 45, 49, 54, 56, 69) (Figure 2). The network graph shows how the available trial evidence was connected. Each node represents either CAU or a peer-support intervention package, defined by the component or combination of components included in that intervention. Several packages included relationship-building (D) and social support (E), suggesting that these components were commonly represented in the three-month quality-of-life evidence network. However, these components usually appeared alongside other components, meaning that the network graph alone does not show which individual component was effective.

**Figure 2.**
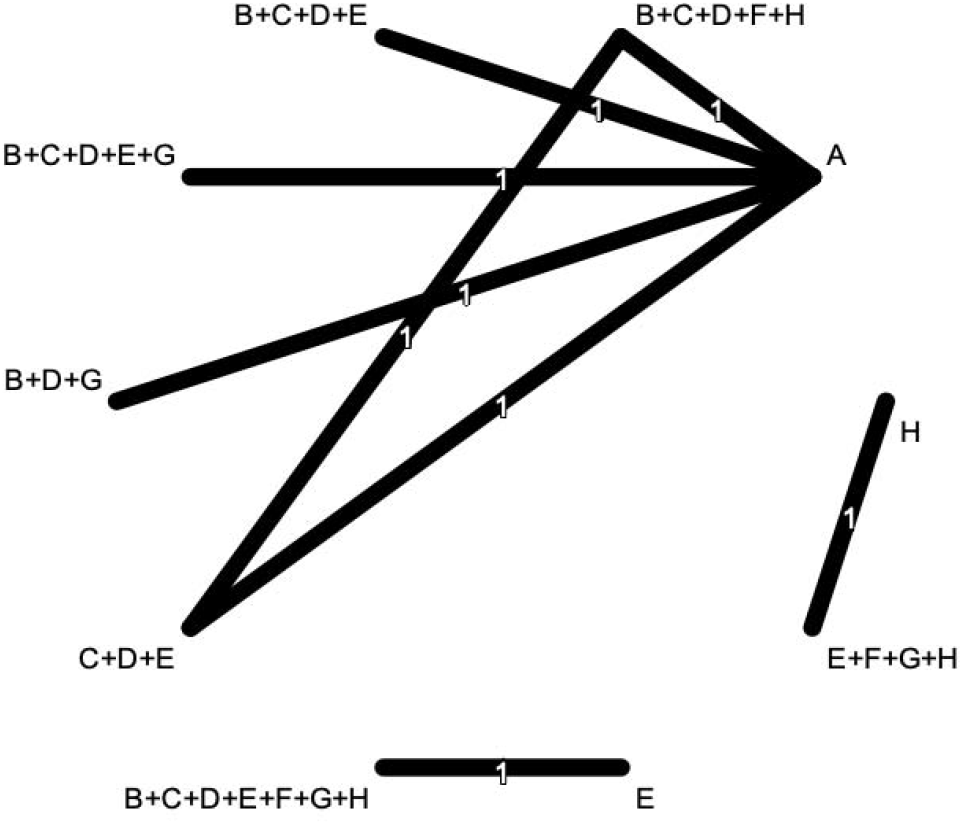
Evidence network for quality of life at three months. A = Comparator arm; B = Training and development; C = Maintaining PSW wellbeing; D = Relationship-building; E = Social support; F = Emotional support; G = Practical support; H = Cultural adaptation. Lines indicate direct comparisons between arms. Numbers on lines indicate the number of studies contributing to each direct comparison; in this figure, all direct comparisons were informed by one trial.

Each direct package comparison was informed by one trial. Because the package network was disconnected, no standard NMA estimate was reported across the full network. The additive CNMA could separately estimate Training and development, Relationship-building and Practical support. Practical support had a positive estimate with a CI excluding zero (incremental SMD=0.52, 95% CI 0.17 to 0.87, *p*=0.003). Estimates for Training and development (-0.06,-0.75 to 0.63) and Relationship-building (-0.39,-1.15 to 0.36) were imprecise and included no effect. Other components were not separately estimable.

#### Quality of life: Six-month follow-up

For quality of life at six months (2,288 participants in 12 studies), the evidence network remained partly disconnected (39, 43, 45, 49, 51, 53–57, 66, 69) (Figure 3). Several packages included multiple peer-support components, indicating that the six-month evidence was also based mainly on multi-component interventions rather than isolated tests of individual components.

**Figure 3.**
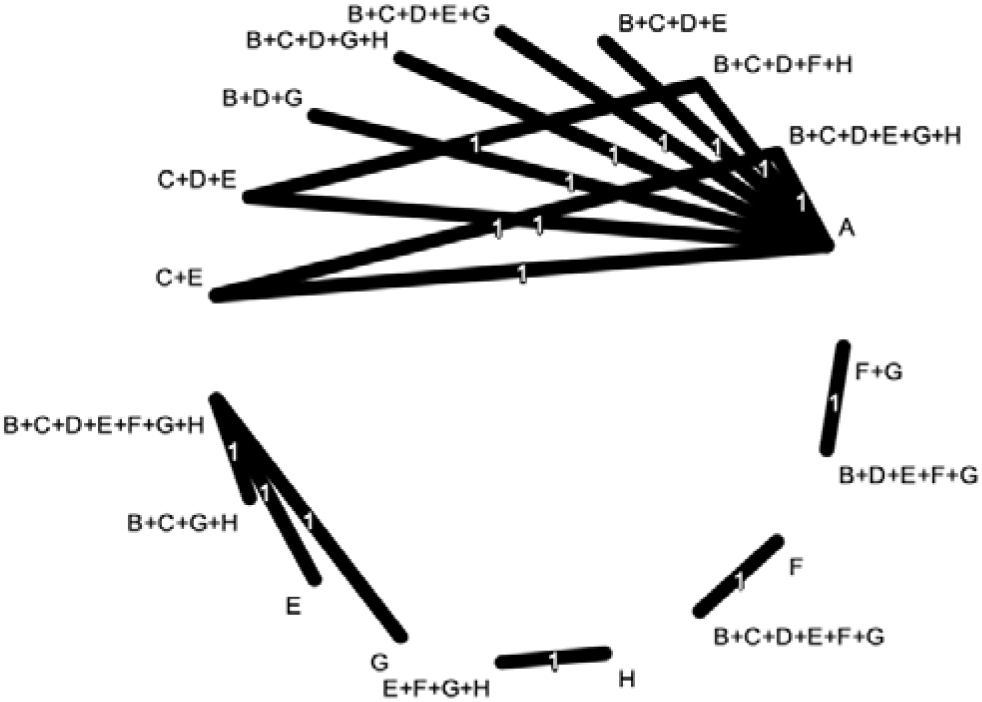
Network graph of QoL at six months. A = Comparator arm; B = Training and development; C = Maintaining PSW wellbeing; D = Relationship-building; E = Social support; F = Emotional support; G = Practical support; H = Cultural adaptation. Lines indicate direct comparisons between arms. Numbers on lines indicate the number of studies contributing to each direct comparison; in this figure, all direct comparisons were informed by one trial.

All seven components were separately estimable at 6 months (Table 5). Estimates ranged from-0.22 to 0.43 and all CIs included zero. Directions were mixed and no component showed a clear or consistent quality-of-life signal at this follow-up.

**Table 5.** Incremental component effects for quality of life at 6 months.

| Component | Incremental SMD | 95% CI | p-value |
| --- | --- | --- | --- |
| B: Training and development | 0.43 | -0.24, 1.11 | 0.21 |
| C: Maintaining PSW wellbeing | -0.22 | -0.71, 0.27 | 0.38 |
| D: Relationship-building | -0.09 | -0.74, 0.56 | 0.78 |
| E: Social support | 0.17 | -0.30, 0.65 | 0.48 |
| F: Emotional support | -0.02 | -0.47, 0.44 | 0.94 |
| G: Practical support | -0.21 | -0.61, 0.20 | 0.31 |
| H: Cultural adaptation | -0.22 | -0.86, 0.42 | 0.50 |

#### Quality of life: 12-month follow-up

Eight trials randomising 1,917 participants contributed quality-of-life data at approximately 12 months (41, 49, 51, 54–56, 66, 69) (Figure 4). The package network was partly disconnected, and most PSW arms combined several components.

**Figure 4.**
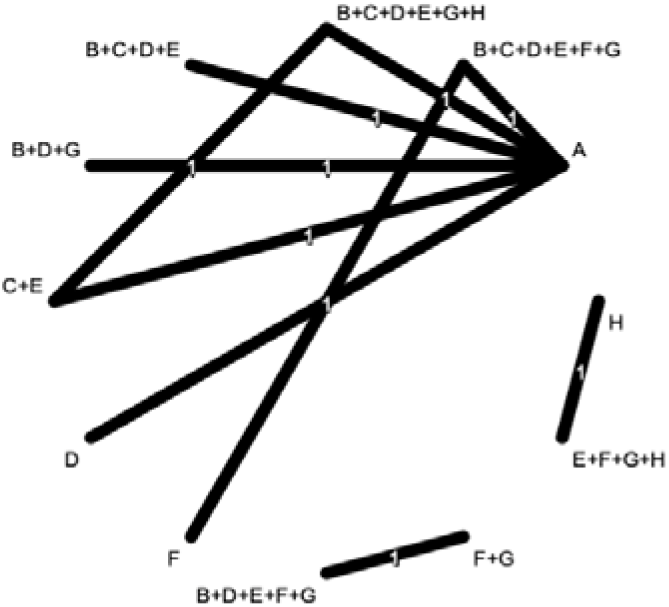
Network Plot for QoL at 12 months. A = Comparator arm; B = Training and development; C = Maintaining PSW wellbeing; D = Relationship-building; E = Social support; F = Emotional support; G = Practical support; H = Cultural adaptation. Lines indicate direct comparisons between arms. Numbers on lines indicate the number of studies contributing to each direct comparison; in this figure, all direct comparisons were informed by one trial.

All seven components were separately estimable (Table 6). Social support had a positive estimate with a CI excluding zero (incremental SMD=1.57, 95% CI 0.12 to 3.01, *p*=0.030), whereas Maintaining PSW wellbeing had a negative estimate (-1.66,-3.05 to-0.28, *p*=0.018). The remaining estimates were imprecise and included no effect. Both isolated estimates had wide CIs and were inconsistent with the corresponding component estimates at earlier follow-ups, limiting causal interpretation.

**Table 6.** Incremental component effects for QoL at 12 months.

| Component | Incremental SMD | CI | p-value |
| --- | --- | --- | --- |
| B: Training and development | 0.47 | -1.49, 2.43 | 0.64 |
| C: Maintaining PSW wellbeing | -1.66 | -3.05, -0.28 | 0.02 |
| D: Relationship-building | 0.24 | -1.24, 1.71 | 0.75 |
| E: Social support | 1.57 | 0.12, 3.01 | 0.03 |
| F: Emotional support | -0.45 | -2.34, 1.43 | 0.64 |
| G: Practical support | -0.54 | -2.01, 0.92 | 0.47 |
| H: Cultural adaptation | -0.25 | -1.92, 1.42 | 0.77 |

#### Recovery: Three-month follow-up

Three studies contributed recovery data at approximately 3 months (49, 50, 54). The evidence comprised three disconnected direct comparisons, each informed by a single trial (Figure 5). Because the intervention network was disconnected and the distribution of components did not allow component effects to be identified, neither a standard NMA across all interventions nor an additive CNMA was presented. Component-level effects were therefore not estimable at this follow-up.

**Figure 5.**
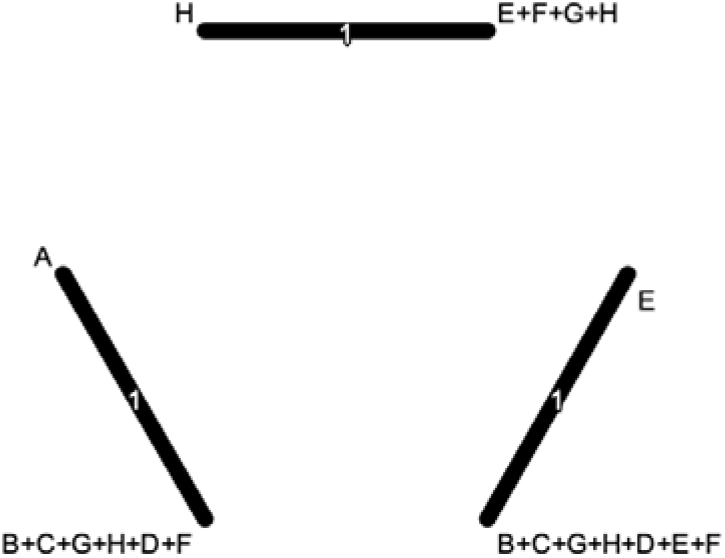
Network Plot for Recovery at three months. A = Comparator arm; B = Training and development; C = Maintaining PSW wellbeing; D = Relationship-building; E = Social support; F = Emotional support; G = Practical support; H = Cultural adaptation. Lines indicate direct comparisons between arms. Numbers on lines indicate the number of studies contributing to each direct comparison; in this figure, all direct comparisons were informed by one trial.

#### Recovery: Six-month follow-up

For recovery at six months (921 participants in six studies (42, 49, 50, 53–55)), Direct package comparisons were sparse and the network was partly disconnected (Figure 6).

**Figure 6.**
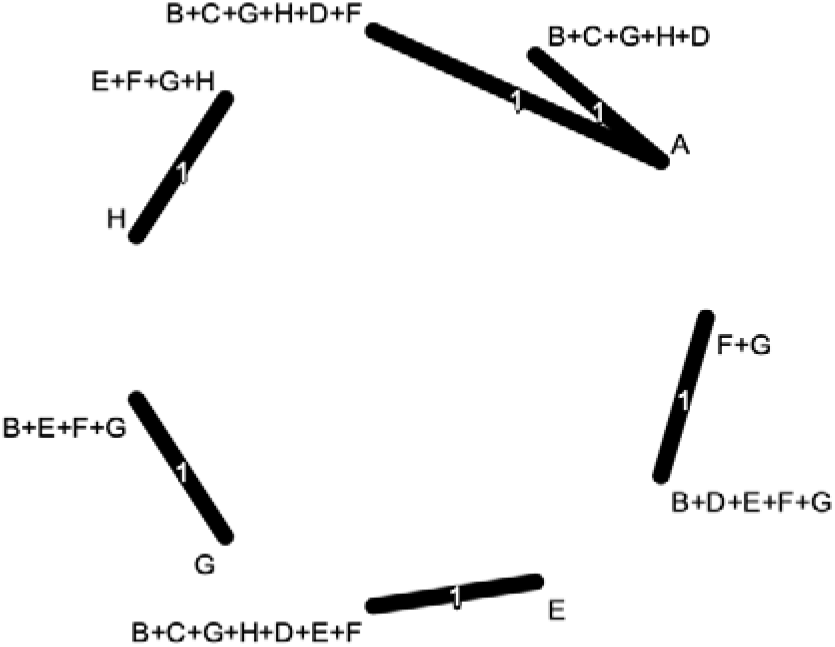
Network Plot for Recovery at six months. A = Comparator arm; B = Training and development; C = Maintaining PSW wellbeing; D = Relationship-building; E = Social support; F = Emotional support; G = Practical support; H = Cultural adaptation. Lines indicate direct comparisons between arms. Numbers on lines indicate the number of studies contributing to each direct comparison; in this figure, all direct comparisons were informed by one trial.

Only Relationship-building and Emotional support were separately estimable. Relationship-building had a positive estimate with a CI excluding zero (incremental SMD=0.90, 95% CI 0.03 to 1.78, *p*=0.042); Emotional support was imprecise and included no effect (-0.10,-0.66 to 0.47).

#### Recovery: 12-month follow-up

Four trials randomising 841 participants contributed recovery data at approximately 12 months (41, 49, 55, 59) (Figure 7). The package network comprised three disconnected sets of comparisons.

**Figure 7.**
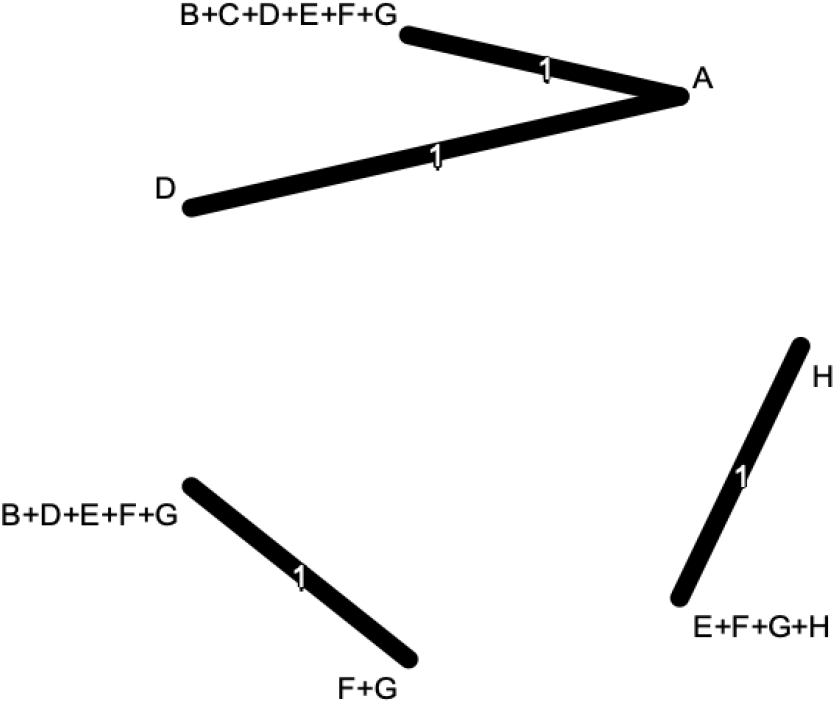
Network Plot for Recovery at 12 months. A = Comparator arm; B = Training and development; C = Maintaining PSW wellbeing; D = Relationship-building; E = Social support; F = Emotional support; G = Practical support; H = Cultural adaptation. Lines indicate direct comparisons between arms. Numbers on lines indicate the number of studies contributing to each direct comparison; in this figure, all direct comparisons were informed by one trial.

Under the additive CNMA model, Relationship-building was the only separately estimable component and had a positive estimate with a CI excluding zero (incremental SMD=0.50, 95% CI 0.29 to 0.72, *p*<0.001; Figure 8). Because the package network was disconnected, this estimate depends on the untestable additivity assumption and should be considered provisional.

**Figure 8.**
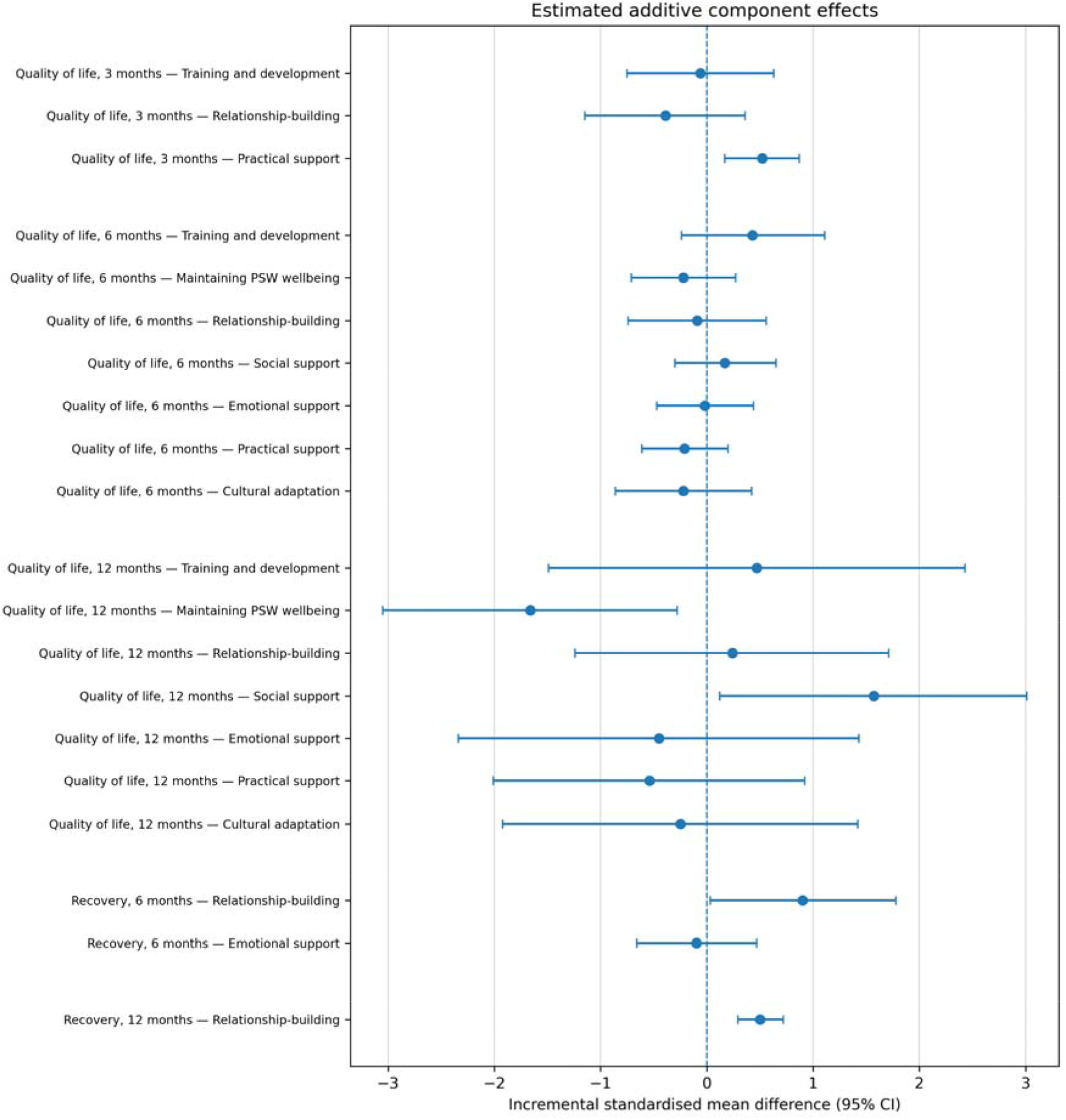
Additive component effect estimates for quality of life and recovery. Positive values favour the component. Estimates are incremental SMDs with 95% CIs; they rely on an additive model, and additivity could not be tested in disconnected networks.

Table 7 summarises the component-level findings and their principal uncertainties.

**Table 7.** Summary of additive component network meta-analysis findings.

| Outcome | Follow-up | Component-level signal | Main uncertainty |
| --- | --- | --- | --- |
| Quality of life | 3 months | Practical support: positive estimate | Only three components estimable; sparse network |
| Quality of life | 6 months | No clear signal | All CIs included no effect; mixed directions |
| Quality of life | 12 months | Social support: positive; Maintaining PSW | Wide CIs; |
|  |  | wellbeing: negative | isolated and inconsistent with earlier follow-ups |
| Recovery | 3 months | Not estimated | Disconnected and too sparse for unique component estimates |
| Recovery | 6 months | Relationship-building: positive estimate | Only Relationship-building and Emotional support estimable |
| Recovery | 12 months | Relationship-building: positive estimate | Disconnected network; additivity assumption untestable |

“No clear signal” indicates that estimated CIs included no effect. “Not estimated” indicates that the component structure did not permit unique estimation. Positive or negative signals are associations under the additive model and should not be interpreted as definitive causal effects.

## DISCUSSION

This component network meta-analysis examined which components of one-to-one PS were associated with outcomes for adults using mental health services. Sufficient data for quantitative synthesis were available for quality of life and recovery. Time-specific component signals were observed for Practical support and Social support in relation to quality of life, while Relationship-building showed the most consistent positive association with recovery. These findings should be interpreted as exploratory because several networks were sparse or partly disconnected, some components were not estimable, estimates were inconsistent across follow-up points, and heterogeneity was high in the 12-month quality-of-life analysis.

For quality of life, practical support was significantly associated with improved outcomes at three months. This finding is clinically plausible. In the early stages of PS, service users may be facing immediate difficulties relating to housing, benefits, medication, appointments, finances, food, or daily functioning (73). Practical support may therefore produce relatively rapid improvements in perceived quality of life by reducing tangible stressors and increasing the person’s sense of safety and control. This is important because PS is sometimes understood primarily as emotional or relational work. Our findings suggest that practical assistance may be an important early mechanism, particularly for people whose mental health difficulties are compounded by social adversity.

At six months, all seven quality-of-life component estimates had confidence intervals that included zero, and the directions of effect were mixed. Training and development had the largest positive point estimate, but there was no clear evidence that this represented a consistent component effect. More broadly, the quality-of-life findings did not show a stable pattern across follow-up points: Practical support was positive at 3 months but not at 6 or 12 months, whereas Social support showed only a small estimate at 6 months before a larger positive estimate at 12 months. Statistical significance may reflect differences in the amount and precision of available evidence rather than genuine changes in component effectiveness over time. These time-specific findings should therefore be interpreted as exploratory rather than as evidence for a sequence in which different components become effective at different stages.

At 12 months, social support was significantly associated with improved quality of life. This suggests that the longer-term value of PS may lie partly in helping service users reconnect with social roles, relationships and community activities (4). Quality of life is broader than symptom reduction; it includes belonging, participation, meaning and everyday functioning (74). Social support may therefore become especially important after the initial period of stabilization, when service users may be ready to rebuild social networks and participate more fully in valued activities (75). This interpretation is consistent with recent arguments that recovery and social inclusion for people with severe mental illness require stronger links between mental health services and civil society, including community organizations, peer networks and other non-clinical resources (76). This finding aligns with the recovery-oriented basis of PS, in which living well with or beyond mental health difficulties is supported through connection, hope and participation.

An unexpected finding was that maintaining PSW wellbeing was negatively associated with quality of life at 12 months. This result should be interpreted with caution. It is unlikely that supporting PSW wellbeing is intrinsically harmful. One possible explanation is that this component was more commonly reported in complex interventions, services working with higher-need populations, or trials with more developed organisational structures and more challenging implementation contexts, where supervision and organisational support are more visible and more extensively described (13). It may also reflect reporting differences: studies that described PSW supervision and wellbeing support in greater detail may have been those with more intensive or complex peer support models (13, 23). The high heterogeneity and wide confidence intervals at 12 months reinforce the need for caution. Nevertheless, this finding is useful because it highlights that organisational support for PSWs should not be assumed to improve service-user outcomes automatically. The content, quality and purpose of supervision and wellbeing support may matter (77). For example, supervision that protects role distinctiveness, supports reflective practice and preserves mutuality may have different effects from supervision that pulls PSWs toward conventional clinical roles.

For recovery, relationship-building was the clearest finding. It was significantly associated with improved recovery at both 6 and 12 months, with a large effect at 6 months and a sustained positive effect at 12 months. This supports the theoretical foundation of PS: the relationship between service user and PSW is not merely a delivery channel but may be a central mechanism of change, consistent with evidence that trusting relationships based on shared lived experience underpin peer worker interventions(78). Relationship-building through mutuality, shared lived experience, trust, goal-setting and appropriate story sharing may help service users reinterpret their experiences, develop hope, and construct a more positive identity beyond illness. Recent realist-informed qualitative evidence similarly suggests that sharing lived experience and role-modelling recovery can facilitate reciprocity, mutuality, self-acceptance, hopefulness and belonging (31). This also aligns with one of the most established frameworks of recovery, the CHIME framework, which emphasizes connectedness, hope, identity, meaning and empowerment (75). The repeated association between relationship-building and recovery strengthens the argument that PS should not be reduced to task-based support or generic case management.

Methodologically, this review suggests that CNMA is useful for examining candidate active ingredients in one-to-one PS, but also highlights the limits of applying this approach retrospectively to relational and context-sensitive psychosocial interventions. Even in a comparatively large PS evidence base, several networks were sparse or partly disconnected, and some components could not be estimated separately. Future research should therefore prospectively specify and manipulate intervention components rather than relying solely on components retrospectively coded from incomplete intervention reports. Large factorial or master-protocol trials may provide a stronger design for disentangling component effects, as illustrated by the RESiLIENT trial, which evaluated multiple cognitive behavioural therapy skills against common controls (79). Future trials should also report intervention content in greater detail, using a shared component typology where possible.

This may be particularly important for Cultural adaptation of PS. Cultural adaptation was the least frequently identified component, but adaptation may have occurred informally or remained unreported. This matters because culturally adapted mental health interventions can produce meaningful benefits (80), and because PS is not culturally neutral. Assumptions about recovery, disclosure, mutuality, help-seeking, independence, family involvement, and social connection may reflect the dominant values of countries where PS has been most widely developed and evaluated (81). Without decentring these assumptions, PS models may be less acceptable or less effective for people from different cultural backgrounds. Future trials should therefore report whether and how PS was culturally adapted, including peer matching, language, family or community involvement, culturally specific meanings of recovery, and norms around support-seeking and disclosure.

This study has several strengths. To our knowledge, it is the first component network meta-analysis of one-to-one PS for adults using mental health services. Its originality lies in moving beyond the question of whether PS works to examine which components, and where possible combinations of components, are associated with benefit. The study was based on a pre-registered protocol, used PRISMA-NMA guidance, updated previous searches (3), incorporated evidence from trial registries and recent umbrella reviews (15, 16), and applied a PS component typology developed through systematised review and expert consultation (32). The analysis therefore provides a transparent synthesis of a complex intervention literature and identifies priorities for intervention design, reporting, implementation, fidelity assessment, and future trial development. Its potential policy and practice relevance is also substantial: the findings highlight the relational core of PS and identify Practical support, Social support and Relationship-building as candidate components for further prospective evaluation, while recognising that these implications are based on preliminary component-level signals.

Several limitations should be acknowledged. First, many networks were sparse, and some outcome domains could not be analysed because of insufficient comparable data. Second, most networks were disconnected chains, limiting the strength of indirect comparisons. Third, heterogeneity was high in some analyses, especially quality of life at 12 months. Fourth, component coding depended on what was reported in trial papers; absence of reporting may not mean absence of a component in practice. Fifth, some components co-occurred frequently, making it difficult to isolate independent effects. Sixth, trials varied in population, setting, comparator, follow-up period, outcome measure and level of organisational support. We did not formally assess selective outcome or time-point reporting, and apparent differences across follow-up periods may therefore partly reflect the availability and precision of reported data. Seventh, the searches were completed in June 2023, approximately three years before submission. The extended interval reflects the substantial work required to code complex interventions, harmonise outcomes and conduct CNMA, but means that more recent eligible trials may not have been captured. Finally, the evidence base remains geographically uneven, with most studies conducted in North America and Europe.

In conclusion, this CNMA provides preliminary evidence that different components of one-to-one PS may be associated with different outcomes over time. Practical support and Social support showed time-specific positive associations with quality of life, while Relationship-building appears particularly relevant to recovery. The findings do not provide definitive prescriptions for PS design, because several networks were sparse, some components were not estimable, estimates were not consistently positive across follow-up points, and intervention reporting was often limited. The findings therefore do not support a prescriptive temporal sequence for PSW practice. Rather, Practical support, Social support and Relationship-building should be considered candidate components for prospective evaluation, with Relationship-building showing the most consistent positive association across the recovery analyses. Future studies should use clearer component reporting, examine mechanisms through prospectively designed trials or mixed-methods process evaluations, and describe cultural adaptation in sufficient detail. Furthermore, integration of the PSW role into the mental health workforce is challenging (82), due to staff attitudes (83), status within the team (84), resource constraints (85), organisational processes (86) and organisational culture (87). Future research will be needed to identify implementation influences on PSW role components. Overall, PS is a relational and context-sensitive intervention; understanding its active ingredients requires methods that can capture both component effects and the contexts in which those components are delivered.

All authors report no conflict of interest.

## Ethics statements

Patient consent for publication: Not applicable. Ethics approval: Not applicable.

## Availability of data and material

All data underlying the findings are provided within the manuscript and its Supporting Information files.

## Supporting information

Supporting Information 1_PRISMA Checklist

## Data Availability

No data was generated by this study. All data underlying the findings are provided within the manuscript and its Supporting Information files.

## Acknowledgement

MS acknowledged the support of the NIHR Nottingham Biomedical Research Centre. AJS/LJG/EAS were supported by the NIHR Applied Research Collaboration East Midlands (ARC EM). The views expressed were those of the authors and not necessarily those of the NIHR or the Department of Health and Social Care.

## Supporting information

**S1 Table. PRISMA Extension for Network Meta-analyses Checklist**

**S2 Table. 26 Studies Identified in Previous Systematised Review to Identify PS Components**

**S3 Table. Search Strategy**

**S4 Table. Searches on Cochrane Central**

**S5 Table. Details of the included studies (n=36)**

**S6 Table. Component coding across the 82 arms of the 36 included studies**

