## Supporting Information 2-6 for "One-to-one peer support work in mental health services: systematic review and component network meta-analysis"

### Supporting Information S2 Table: 26 Studies Identified in Previous Systematised Review to Identify PS Components

### Supporting Information S3 Table: Search Strategy

| # | **Searches** |
| --- | --- |
| 1 | exp peer group/ |
| 2 | "peer* support*".mp. |
| 3 | (peer* adj4 support*).mp. |
| 4 | (peer adj4 (provid* or consumer* or survivor* or specialist* or companion* or recovery or coach* or influenc*)).mp. |
| 5 | (peer* adj4 (tutor* or mentor* or educat* or intervention* or listen* or mediat* or befriend* or therap* or worker* or counsel*)).mp. |
| 6 | ("mental health peer*" or "lived experience*").mp. |
| 7 | ("one to one" or "one on one" or "1 to 1" or intentional or individual or "peer to peer").mp. |
| 8 | 1-to-1.mp. |
| 9 | exp mental health/ |
| 10 | exp mental health service/ |
| 11 | (mental adj4 (health or disease* or illness* or disorder*)).mp. |
| 12 | (depress* or "manic episode*" or mania or schizophren* or bipolar or delusion* or "personality disorder*" or stress or "obsessive compulsive" or neurotic or phibi* or agrophobi*).mp. |
| 13 | (psychiatr* or "severe mental" or psychopath* or "drug addict*" or "substance abuse" or "substance misuse" or ptsd).mp. |
| 14 | mental well being.mp. |
| 15 | ("dissociative identity disorder*" or "dissociative amnesia" or "disocciative dosorder*" or phobi* or "body dysmorphi*" or hoard* or "gender identit*" or anorexi* or bulimi* or purg* or bing* or eating disorder*).mp. |
| 16 | 1 or 2 or 3 or 4 or 5 or 6 |
| 17 | 7 or 8 |
| 18 | 9 or 10 or 11 or 12 or 13 or 14 or 15 |
| 19 | 16 and 17 and 18 |
| 20 | randomized controlled trial.pt. |
| 21 | controlled clinical trial.pt. |
| 22 | randomized.ab. |
| 23 | placebo.ab. |
| 24 | clinical trials as topic.sh. |
| 25 | randomly.ab. |
| 26 | trial.ti. |
| 27 | 20 or 21 or 22 or 23 or 24 or 25 or 26 |
| 28 | exp animals/ not humans.sh. |
| 29 | 27 not 28 |
| 30 | 19 and 29 |

The search strategy for Embase was developed using Cochrane guidance^1 2^. The Embase OVID RCT search filter was used.

1. The Cochrane Collaboration. Embase syntax: a comparison with Ovid. London: Author, 2012.

2. Glanville J, Foxlee R, Wisniewski S, et al. Translating the Cochrane EMBASE RCT filter from the Ovid interface to Embase.com: a case study. *Health Info Libr J* 2019;36(3):264-77. doi: 10.1111/hir.12269

### Supporting Information S4 Table: Searches on Cochrane Central

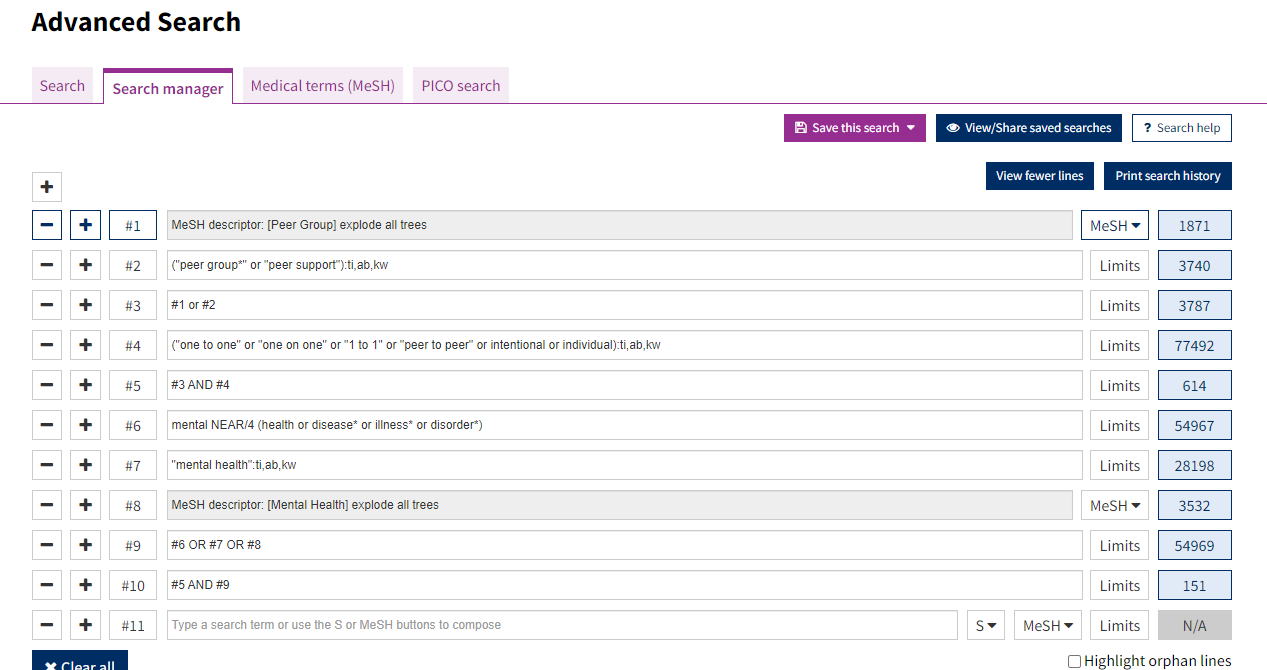

### Supporting Information S5 Table: Details of the included studies (n=36)

| **No** | **Study** | **Country** | **Population** | **Intervention** | **Control** | **Intervention** | **Control** | **Outcomes** | **Assessments** | **Longest Follow Up** | **Role of PS** | **Organisational Support** |
| --- | --- | --- | --- | --- | --- | --- | --- | --- | --- | --- | --- | --- |
| #01 | Gillard et al. 2022 | England | Adult psychiatric inpatients (age ≥18 years) with at least one previous admission in the preceding 2 years, excluding those who had a diagnosis of any organic mental disorder, or a primary diagnosis of learning disability, an eating disorder, or drug or alcohol dependency, recruited from seven state-funded mental health services in England | 294 | 296 (available data 306:267:3) | Manual-based, one-to-one peer support, focused on building individual strengths and engaging with activities in the community plus CAU | CAU - community mental health services | 1) Subjective quality of life 2) Social inclusion 3) Hope 4) Psychiatric symptom | 1) Manchester Short Assessment of Quality of Life 2) Objective Social Outcomes Index 3) Herth Hope Index 4) Brief Psychiatric Rating Scale | 12 months | A | H |
| #02 | Rohrbach et al. 2022 | Netherlands | Adults with eating disorders | 90 (89:1) Featback (internet intervention) plus weekly PS  87 (84:3) Weekly PS only | 90 (88:2) Waitlist  88 (82:6) Featback (internet intervention) only | PSW used their own experience to help others with eating disorder symptoms via chat and email for 20 minutes per week for 8 weeks. | Waitlist control, and internet only | 1) Eating disorder symptomatology 2) Anxiety and depression 3) General self-efficacy 4) Experienced social support 5) Motivation, satisfaction, and help-seeking 6) Similarity of oneself to PSW | 1) Eating Disorder Examination Questionnaire 2) Patient Health Questionnaire-4 3) General Self-Efficacy Scale 4) Social Support List 5) Self-made questionnaire 6) Inclusion of Other in the Self scale at Week 3 | 12 months | A for Featback+PS  S for PS only | H |
| #03 | Tinland et al., 2020 | France | Adults with a DSM-5 diagnosis of schizophrenia, bipolar I disorder, or schizoaffective disorder | 196 (69:127) | 198 (86:112)   Total 155: 239 | Peer support workers with a readmission reduction tool | CAU | 1) Therapeutic alliance 2) Quality of life 3) Mental health symptoms 4) Empowerment 5) Recovery outcomes | 1) Alliance Scale 2) Schizophrenia Quality-of-Life scale 3) Colorado Symptom Index 4) Empowerment Scale 5) Recovery Assessment Scale | 6 months | A | H |
| #0$ | Kidd et al. 2021 | Canada | Adults with schizophrenia spectrum illnesses in the period immediately following hospitalization. | 41 (19:22) | 22 (6:16) Brief intervention.  44 (16:28) CAU | Provision of a modest amount of personalised supplies given to assist in the transition of discharge, and help with key tasks of community living | Brief intervention, and CAU | 1) Community ability 2) Community engagement 3) Symptomatology 4) Substance use 5) Personal recovery 6) Social support 7) Quality of life | 1) Multnomah Community Ability Scale 2) Community Integration Scale 3) Brief Symptom Inventory 4) SubstanceDisorder subscale of the GAIN-Short Screener 5) Personal Recovery Outcome Measure 6) Social Support Survey 7) Satisfaction with Life scale | 6 months | A | H |
| #05 | Maru et al. 2021 | USA | Adults with psychiatric disabilities receiving outpatient community-based mental health services | 83 (43:40) | 83 (42:41) | Clients and peer support workers meet as frequently as needed, on a weekly or biweekly basis, through one-on-one in-person or phone meetings. Peer support workers support clients choosing meaningful vocational options through the identification of personal preferences. | Peer support CAU | 1) Vocational and prevocational activity 2) Quality of life (QOL) 3) Hope 4) Work readiness 5) Working alliance | 1) Vocational and prevocational activity scale 2) Lehman’s Brief Quality of Life Interview 3) The Work Hope Scale 4) The Work Readiness Scale 5) Working Alliance Inventory-Short Form | 12 months | A | H |
| #06 | Byrne et al., 2020 | USA | Adults hospitalised with Substance Use Disorder (SUD) complications | 51 (19:32) | 47 (21:26) | Recovery coaching offered on average 1 to 2 sessions weekly | CAU | 1) Substance use frequency 2) Physical and mental helath | 1) Addiction Severity Index 2) The 12-item Short Form Health Survey | 6 months | A | H |
| #07 | Ranzenhofer et al., 2020 | USA | Outpatients with anorexia nervosa, bulimia nervosa, or binge-eating disorder. | 20 (20:0) | 18 (17:1)  22 (22:0) | Peer mentors act as a support person and role model, providing interpersonal support and guidance based on training and personal experiences. | Social support mentorship by a person without experience of an eating disorder | 1) Eating disorder symptoms 2) Eating disorder QOL 3) Anxiety symptoms 4) Depression symptoms | 1) Eating Pathology Symptoms Inventory 2) Eating Disorder Quality of Life assessment survey 3) State-Trait Anxiety Inventory 4) Patient Health Questionnaire-9 | 6 months | A | H |
| #08 | Cardi et al., 2019 | England | Adult with anorexia nervosa | 99 (96:3) | 88 (84:4) | Having access a workbook, a library of short video clips (‘vodcasts’) and six 1-hour text-chat sessions with PSW for 6 weeks. PSW worked based on the cognitive interpersonal model and recovery framework, called RecoveryMANTRA (Maudsley Model of Anorexia Nervosa Treatment for Adults). | CAU | 1) Eatind disorder symptoms 2) Mood 3) Quality of life 4) Motivation for treatment 5) Alliance with the therapist and cognitive and behavioural flexibility | 1) Eating Disorder Examination Questionnaire 2) Depression, Anxiety and Stress Scales 3) Work and Social Adjustment Scale 4) Autonomous and Controlled Motivations for Treatment Questionnaire 5) Self-made questionnaires | 12 months | A | H |
| #09 | Pfeiffer et al., 2019 | USA | Adult psychiatric inpatients at high risk for suicide | 34 | 36  Total 37:29:4 | Peer support using a suicide prevention tool plus CAU | CAU | 1) Suicide attempts 2) Suicidal ideation 3) Hopelessness 4) Hope 5) Belongingness 6) Use of services | 1) Columbia Suicide Severity Rating Scale 2) Beck Scale for Suicidal Ideation 3) Beck Hopelessness Scale 4) Hope Scale 5) NIH Adult Toolbox Social Relationship scales 6) Checklist including inpatient, emergency room, intensive outpatient, outpatient, and community support/self-help services (Booth, Kirchner, Fortney, Ross, & Rost, 2000). | 6 months | A | H |
| #10 | Shorey et al., 2019 | Singapore | Mothers at risk of postnatal depression | 69 (69:0) | 69 (69:0) | Technology-based peer-support intervention program | CAU | 1) Postnatal depression 2) Postnatal anxiety 3) Loneliness 4) Perceived social support | 1) Edinburgh Postnatal Depression Scale, and Patient Health Questionnaire 2) State-Trait Anxiety Inventory 3) University of California, Los Angeles Loneliness Scale 4) Perceived Social Support for Parenting | 3 months | A | H |
| #11 | Corrigan et al., 2018 | USA | Adult Latinos with serious mental illness | 55 (28:26:1) | 55 (36:18:1) | PSW used cognitive-rehabilitation and social skills in face-to-face meetings at least once per week for 6 months. | CAU | 1) Recovery 2) Personal empowerment 3) Quality of life | 1) Recovery Assessment Scale 2) Righteous Anger subscale of the Empowerment Scale 3) Lehman’s Quality of Life Scale | 12 months | A | L |
| #12 | Johnson et al., 2018 | England | Adult patients currently on the caseload of crisis resolution teams for at least a week because of a psychiatric crisis | 220 (132:88) | 219 (131:87) | Peer-supported self-management plus CAU | CAU – community mental health services plus self-management workbook | 1) Overall psychiatric symptoms 2) Social network support 3) Recovery 4) Satisfaction with services 5) Hospitalisation | 1) Brief Psychiatric Rating Scale 2) Lubben Social Network Scale 3.a) Illness Management & Recovery Scale (patient version) 3.b) Questionnaire on the Process of Recovery 4) Client Satisfaction Questionnaire 5) Community tenure (days) | 18 months | A | H |
| #13 | Mahlke et al., 2017 | Germany | Patients, aged 18–80, using in- and out-patient services with primary diagnosis of schizophrenia and related disorders, affective disorders, or personality disorder and a duration of illness of more than 2 years. | 114 (65:49) | 112 (59:53) | Community-based peer support for individual recovery plus CAU | CAU - in-patient and out-patient mental health as usual | 1) Overall psychiatric symptoms 2) Quality of Life 3) Social functioning 4) Empowerment 5) Hospitalisation | 1) Clinical Global Impression: Severity scale 2) Modular System for Quality of Life and EuroQol Questionnaire EQ. 5D 3) Global Assessment of Functioning 4) General Self-Efficacy Scale 5) Days in hospital | 12 months | A | H |
| #14 | Seeley et al., 2017 | USA | Patients, aged 55 and above, referred to an intergovernmental agency and meeting criteria for mild to moderate depression and/or anxiety | 31 | 31   Total 50:12) | Peer-supported cognitive behavioural intervention for mild-moderate depression and/ or anxiety | Waitlist control | 1) Depression¥ 2) Anxiety 3) Working Alliance | 1) PHQ-9 2) GAD-7 3) Working Alliance Inventory | 2.5 months | A | L |
| #15 | Yamaguchi et al., 2017 | Japan | Patients, age 20 years or older, using outpatient psychiatric clinic or psychiatric hospital in Tokyo, who received services from case managers in either a psychiatric day care or visiting nurse program. | 26 (10:16) | 27(12:15) | Peer supported shared decision-making plus CAU | CAU - medical consultation | 1) Overall psychiatric symptoms 2) Social Functioning 3) Empowerment 4) Working Alliance | 1) The Brief Psychiatric Rating Scale 2) Global Assessment of Functioning (GAF) 3) Patient Activation Measure 4) Scale To Assess Therapeutic Relationships in Community Mental Health Care (STAR) – Clinician & Patient versions | 12 months | A | L |
| #16 | Rogers et al., 2016 | USA | Clients, over the age of 18, who were court ordered for treatment because of a psychiatric crisis civilly committed for a mental health crisis, adjudicated by the state court to meet the definition of “a person with a serious mental illness,” | 63 (34:29) | 50 (29:21) | Individual peer-supported social inclusion and recovery support plus CAU | CAU - Peer-provided services (excluding individual peer support; e.g. social activities, educational courses, group peer support) | 1) Social network support 2) Overall psychiatric symptoms 3) Recovery 4) Quality of Life | 1) Interpersonal Support Evaluation List 2) BASIS-R 3) Recovery Assessment Scale 4) Brief Quality of Life | 6 months | A | H |
| #17 | Salzer et al., 2016 | USA | Patients, aged 18 and above, using community outpatient mental health programmes with a diagnosis on the schizophrenia spectrum, bipolar disorder, or major depression | 50 (23:26:1) | 49 (26:23:0) | Peer-delivered support for independent living plus CAU | CAU - usual outpatient mental health care | 1) Quality of life 2) Recovery 3) Empowerment 4) Working Alliance | 1) Lehman’s Quality of Life Interview 2) Recovery Assessment Scale 3) The Empowerment Scale 4) Working Alliance Inventory | 12 months | A | H |
| #18 | Valenstein et al., 2016 | USA | Veterans (adults) with clinical diagnosis of depression, at least one prior antidepressant or psychotherapy trial, and significant depressive symptoms or significant functional limitations. | 144 (27:117) | 243 (47:196) | Telephone-delivered mutual PS, focusing on focused on communication skills, behavioural activation, goal setting, and self-management. Less structured than other PS programmes. | CAU with written self-help materials | 1) Depression 2) Mental health and physical health 3) Quality of life 4) Suicidal thoughts 5) Mental health recovery | 1) Beck Depression Inventory Second Edition 2) Veterans RAND 36-Item Health Survey 3) Quality of Life Enjoyment and Satisfaction Questionnaire Short Form 4) Beck Scale for Suicide Ideation 5) Mental Health Recovery Measure | 12 months | A | H |
| #19 | Wrobleski et al., 2015 | Canada | Adult patients receiving care from a community mental health service with a persistent mental illness, that is significantly affecting daily functioning or a person with both a mental health diagnosis and substance use issue | 12 (11:1) | 9 (7:2) | Peer-supported self-management plus occupational therapy | Self-management support from a (non-peer) mental health worker plus occupational therapy | 1) Quality of Life | 1) Lehman’s Quality of Life Interview | 6 months | S | H |
| #20 | Simpson et al., 2014 | England | Inpatients, aged 18–65, approaching discharge/extended leave from acute mental health inpatient unit | 23 (7:16) | 23 (3:20) | Peer support plus CAU | CAU - community mental health services | 1) Hope 2) Quality of Life 3) Hospitalisation | 1) Beck Hopelessness Scale 2) EuroQol (EQ-5D) 3) Hospitalised | 3 months | A | H |
| #21 | Chinman et al., 2013 | USA | Current adult VA intensive case management patients who have had 30 psychiatric inpatient days or 3 psychiatric admissions in the past year with an Axis 1 psychiatric disorder. | 122 (12:110) | 116 (16:100) | Floating, additional peer-supported case management plus CAU | CAU - case management from community-based Intensive Case Management services | 1) Quality of Life 2) Recovery 3) Empowerment 4) Overall psychiatric symptoms | 1) Lehman’s Quality of Life Interview 2) The Mental Health Recovery Measure Illness Management and Recovery Scale 3) Patient Activation Measure 4)BASIS-R | QoL - 6 months Other - 12 months | A | H |
| #22 | Gjerdingen et al., 2013 | USA | Mothers aged 16 years and older, with a 0- to 6-month-old infant, and depressive symptoms | 13 (13:0) | 14 (14:0) Duola group 12 (12:0) | Telephone PS over 3 months, where timing and frequency of calls tailored to the preferences of the mothers. | CAU | 1) Depression and anxety 2) Depression 3) Overall health status 4) Available Support 5) Importance of Support 6) Satisfaction with Support 7) Illness Days | 1) PHQ-9 2) Center for Epidemiologic Studies Depression Scale (CES-D) 3) A single-item global health measure taken from the EQ-5D4)  4) Available Support 5) Importance of Support 6) Satisfaction with Support 7) Illness Days | 6 months postenrollment | A | H |
| #23 | Proudfoot et al., 2012 | Australia | Adults diagnosed with bipolar disorder by a health professional within the past 12 months and currently being treated | 134 (98:36) | 139 (93:46) Psycho-education only.  134 (93:41) Brief online texts about bipolar disorder were sent. | Online peer coaching plus online psycho-education programme. | 1) Online psycho-education programme 2) attention control | 1) Depression and anxiety 2) Social functioning 3) Empowerment | 1) Goldberg Anxiety and Depression Scale 2) Work and Social Adjustment Scale 3) Multi-dimensional Health Locus of Control | 6 months | A | L |
| #24 | Letourneau et al., 2011 | Canada | Mothers with Postpartum depression, caring for a healthy singleton less than 9 months of age | 27 (27:0) | 33 (33:0) | Home-based PS included maternal-infant interaction teaching for 12 weeks (9 visits on average, duration of visit was 20 min or longer) | Waitlist control | 1) Maternal-infant interactions 2) Depression 3) Social support 4) Infant development 5) Salivary cortisol | 1) Nursing child assessment satellite training 2) Edinburgh Postnatal Depression Scale (EPDS)  3) Social Provisions Scale 4) Bayley Mental Development Index 5) ELISA kit | 12 weeks | A | H |
| #25 | Simon et al., 2011 | USA | Participants, aged 19 or over, who were currently in treatment for bipolar disorder | 64 | 54  Total 85:33 | Online peer recovery coaching plus online recovery planning | Online recovery planning | 1) Engagement with services | 1) Use of online program components - engagement with recovery plans, use of social networking features, use of self-monitoring tools. | 3 weeks | A | H |
| #26 | [Sledge](https://bmcpsychiatry.biomedcentral.com/articles/10.1186/s12888-020-02923-3#ref-CR45) et al., 2011 | USA | Adult inpatients who have experienced three or more psychiatric hospitalizations (or two admissions plus more than three psychiatric ED visits) during the 18-month period prior to recruitment and have a documented diagnosis of schizophrenia, schizoaffective disorder, psychotic disorder not otherwise specified, bipolar disorder or major depressive disorder with or without psychotic features | 39 (22:17) | 37 (16:21) | Community-based peer recovery mentor plus CAU | CAU - community mental health care | 1) Hospitalisation 2) Overall psychiatric symptoms 3) Social Functioning 4) Hope 5) Satisfaction with services 6) Social network support 7) Wellbeing | 1) Number of readmissions - Days in hospital, Hospitalised and Community tenure 2) Brief Psychiatric Rating Scale 3) The Social Functioning Scale 4) The Dispositional Hope Scale 5) Mental Health Statistics Improvement Programme (MHSIP) 6) Sense of Community Index 7) 36 item Short Form Health Survey | 9 months | A | H |
| #27 | Dennis et al., 2009 | Canada | Women in the first two weeks postpartum identified as high risk for postnatal depression | 349 (349:0) | 352 (352:0) | Proactive telephone-based PS for 12 weeks. | CAU | 1) Postnatal depression 2) Anxiety 3) Loneliness 4) Health service utilisation | 1) Edinburgh Postnatal Depression Scale (EPDS) with a structured clinical interview for depression (SCID) 2) State-Trait Anxiety Inventory 3) Short version UCLA Loneliness Scale 4) "bmj.com" p.281 | 24 weeks | A | H |
| #28 | [Rivera](https://bmcpsychiatry.biomedcentral.com/articles/10.1186/s12888-020-02923-3#ref-CR42) et al., 2007 | USA | Adults recruited from inpatient units at a city hospital whom have a diagnosis of a psychotic or mood disorder on axis I, and have had two or more psychiatric hospitalizations in previous two years | 70 (35:35) | 66 (31:35) CAU  67 (34:33) Clinic-based care by psychologist and social worker | Consumer-assisted intensive case management | 1) Intensive case managementa 2) Standard case management (i.e. office-based without intensive components) | 1) Overall psychiatric symptoms 2) Quality of Life 3) Social network support 4) Wellbeing 5) Hospitalisation | 1) Brief Symptom Inventory 2) Lehman Quality of Life Inventory 3) Modification of Pattison Network Inventory  4) Lehman’s Quality of Life - health subscale  5) Days in hospital (per month) | 12 months | A | L |
| #29 | [Sells](https://bmcpsychiatry.biomedcentral.com/articles/10.1186/s12888-020-02923-3#ref-CR39) et al., 2006 | USA | Adult patients currently using local mental health authorities with a primary diagnosis of SMI (schizophrenia spectrum disorder, major mood disorder, or both) and treatment disengagement | 68 | 69 (in total 53:84) | Peer-based case management from peer mental health service provider | Case management as usual from assertive community treatment teams | 1) Working alliance - client 2) Engagement with services | 1) Barrett-Lennard Relationship Inventory (BLRI) modified version 2) Level of Care Utilization System | 12 months | A | L |
| #30 | [Craig](https://bmcpsychiatry.biomedcentral.com/articles/10.1186/s12888-020-02923-3#ref-CR35) et al., 2004 | England | Adult service users currently registered with assertive outreach team and have SMI, with a record of poor engagement, multiple hospitalisations and a high prevalence of problematical behaviours and substance abuse. | 24 | 21 (in total 15:30) | Consumer Health Care Assistant plus CAU | CAU - case management from Assertive Outreach Team | 1) Social functioning 2) Social network support 3) Hospitalisation 4) Satisfaction with services 5) Service engagement | 1) Life Skills Profile 2) Significant others scale 3) Days in hospital and Hospitalised 4) Verona Service Satisfaction Scale (VSSS) 5) Number of missed (DNA) appointments with services. | 12 months | A | H |
| #31 | Davidson et al., 2004 | USA | People with a serious mental illness (such as mood, anxiety, or schizophrenia-spectrum disorders) while being psychiatrically stable for the past six months (i.e., not hospitalised or institutionalised during this period) | 95 PS with $28 stipend | 70 in passive control with $28 stipend  95 in Partnered by nonconsumer with $28 stipend. Total gender split 260 (148: 112) | PS to engage in social or recreational activities together in the community for 9 months, with the expectation of spending 2-4hr/week together. | 1) Stipend 2) Stipend and nonconsumer partner | 1) Psychiatric symptoms 2) Functioning impairment 3) Self-esteem 4) Social functioning 5) Depression 6) Wellbeing 7) Nonpsychotic Psychiatric Symptoms 8) Satisfaction with service | 1) Brief Psychiatric Rating Scale 2) Global Assessment of Functioning 3) Rosenberg Self-esteem Scale 4) Social Functioning Scale 5) Center for Epidemiological Studies—Depression Scale 6) Well Being Scale 7) Global Health Questionnaire 8) Satisfaction with Service | 9 months | A | H |
| #32 | Dennis et al., 2003 | Canada | Postpartum depression mothers between 8 and 12 weeks postpartum, aged at least 18 years, had a singleton birth at 37 weeks’ gestation or more | 20 (20:0) | 22 (22:0) | Telephone PS, where contact frequency was decided by participants | CAU | 1) Depression 2) Self-esteem 3) Child-care stress 4) Loneliness 5) Perceptions of peer support | 1) Edinburgh Postnatal Depression Scale 2) Rosenberg Self-Esteem Scale 3) Child-Care Stress Checklist 4) UCLA Loneliness Scale 10 5) Peer Support Evaluation Inventory | 8 weeks | A | L |
| #33 | Clarke et al., 2000 | USA | Adult patients with a severe mental disorder, a schizophrenic, major affective, or paranoid disorder, or another severe mental disorder, and a documented history of persistent psychotic symptoms other than those caused by substance abuse. | 57 (21:36) | 57 (22:35) Non-peer assertive community treatment  49 (21:28) CAU | Consumer-staffed assertive community treatment | 1) Non-consumer assertive community treatment 2) CAU – usual community mental health services | 1) Hospitalisation | 1) Hospitalised and Community tenure (days) | 6 months | S | L |
| #34 | Hunkeler et al., 2000 | USA | Adults primary care patients with a diagnosis of major depressive disorder or dysthymia and given a prescription of for a SSRI antidepressant (fluoxetine hydrochloride or paroxetine) | 62 Telehealth care and PS | 117 Telehealth care  123 (in total 208:94) CAU | Peer support via telephone contact or face-to-face plus nurse telehealth care plus nurse telehealth plus CAU | 1) Nurse telehealth care plus CAU 2) CAU – usual physician care | 1) Depression and anxiety 2) Social functioning 3) Satisfaction with services | 1) Hamilton Depression Rating Scale- self report version and Beck depression Inventory 2) SF-12 Mental and Physical Composite Scales 3) Patient satisfaction with treatment scale | 6 months | A | L |
| #35 | [Klein](https://bmcpsychiatry.biomedcentral.com/articles/10.1186/s12888-020-02923-3#ref-CR28) et al., 1998 | USA | Adult patients receiving intensive care management with dual diagnosis who had been in community care at the mental health centre for 1 year | 10 | 51 (in total 16:45) | Peer-supported community enablement plus CAU | CAU - Intensive Case Management | 1) Hospitalisation 2) Social functioning 3) Quality of Life 4) Social network support 5) Wellbeing | 1) Days in hospital 2) Global Assessment of Functioning (GAF) 3) Lehman’s Quality of Life (QOL)  4) Lehman’s Quality of Life – Friends subscale 5) Lehman’s Quality of Life – Health subscale | 6 months | A | H |
| #36 | Solomon et al., 1995 | USA | Adults currently on community mental health centre caseload who meet all three criteria for intensive case management and were identified to be at risk for hospitalisation with a diagnosis of major mental illness and a significant treatment history | 46 (27:19) | 45 (16:29) | Consumer case management | Case management as usual from community mental health services | 1) Overall psychiatric symptoms 2) Social network support 3) Quality of life 4) Hospitalisation 5) Working Alliance | 1) Brief Psychiatric Rating Scale (BPRS) 2) Pattison’s Social Network 3) Lehman’s Quality of Life Interview 4) Days in hospital 5) Working Alliance Inventory - staff and client | 24 months | S | H |

PS=Peer support, A=Adjunctive, S=Substitutive, L=Low level of organisational support for peer support (including no reporting of organisational support), H=High level of support for peer support.

1. The Cochrane Collaboration. Embase syntax: a comparison with Ovid. London: Author, 2012.

2. Glanville J, Foxlee R, Wisniewski S, et al. Translating the Cochrane EMBASE RCT filter from the Ovid interface to Embase.com: a case study. *Health Info Libr J* 2019;36(3):264-77. doi: 10.1111/hir.12269

### Supporting Information S6 Table: Component coding across the 82 arms of the 36 included studies

| **No** | **Arm** | **B. Training and development** | **C. Maintaining PSW wellbeing** | **D. Relationship-building** | **E. Social support** | **F. Emotional support** | **G. Practical support** | **H. Cultural adaptation** |
| --- | --- | --- | --- | --- | --- | --- | --- | --- |
| #01 | PS | ✓ | ✓ | ✓ | ✓ |  | ✓ |  |
| #01 | Non-PS |  |  |  |  |  |  |  |
| #02 | Non-PS: Online self-help |  |  |  |  | ✓ |  | ✓ |
| #02 | PS: Chat/email with PSW | ✓ | ✓ |  |  |  |  |  |
| #02 | PS: Online self-help with PSW | ✓ | ✓ |  |  | ✓ |  | ✓ |
| #02 | Non-PS: Waitlist |  |  |  |  |  |  |  |
| #03 | PS |  |  | ✓ |  |  |  |  |
| #03 | Non-PS |  |  |  |  |  |  |  |
| #04 | PS: 10 hours | ✓ |  |  | ✓ | ✓ | ✓ |  |
| #04 | PS: 4 hours | ✓ |  |  | ✓ | ✓ | ✓ |  |
| #04 | Non-PS: Practical support |  |  |  |  |  | ✓ |  |
| #05 | PS: PS by vocational support expert | ✓ | ✓ | ✓ | ✓ | ✓ | ✓ | ✓ |
| #05 | PS: PS by non-vocational support expert | ✓ | ✓ |  |  |  | ✓ | ✓ |
| #06 | PS | ✓ |  | ✓ | ✓ | ✓ | ✓ |  |
| #06 | Non-PS |  |  |  | ✓ |  | ✓ |  |
| #07 | PS | ✓ | ✓ | ✓ |  | ✓ |  | ✓ |
| #07 | Non-PS: Social support |  | ✓ | ✓ | ✓ |  |  |  |
| #07 | Non-PS: Waitlist |  |  |  |  |  |  |  |
| #08 | PS | ✓ | ✓ | ✓ | ✓ | ✓ |  |  |
| #08 | Non-PS |  |  |  |  |  |  |  |
| #09 | PS | ✓ | ✓ | ✓ | ✓ | ✓ | ✓ |  |
| #09 | Non-PS |  |  |  |  | ✓ |  |  |
| #10 | PS | ✓ |  | ✓ | ✓ | ✓ |  |  |
| #10 | Non-PS |  |  |  |  |  |  |  |
| #11 | PS |  |  |  | ✓ | ✓ | ✓ | ✓ |
| #11 | Non-PS |  |  |  |  |  |  | ✓ |
| #12 | PS | ✓ | ✓ | ✓ |  | ✓ | ✓ | ✓ |
| #12 | Non-PS: Recovery workbook |  |  |  |  | ✓ |  |  |
| #13 | PS | ✓ | ✓ | ✓ | ✓ | ✓ | ✓ |  |
| #13 | Non-PS: Psychological treatment |  |  |  |  | ✓ |  |  |
| #14 | PS | ✓ | ✓ | ✓ |  | ✓ | ✓ |  |
| #14 | Non-PS: Waitlist |  |  |  |  |  |  |  |
| #15 | PS | ✓ | ✓ | ✓ |  |  | ✓ | ✓ |
| #15 | Non-PS |  |  |  |  |  |  |  |
| #16 | PS | ✓ | ✓ | ✓ | ✓ | ✓ | ✓ | ✓ |
| #16 | Non-PS: Group education activities |  |  |  | ✓ |  |  |  |
| #17 | PS | ✓ |  | ✓ | ✓ | ✓ | ✓ |  |
| #17 | Non-PS |  |  |  |  | ✓ | ✓ |  |
| #18 | PS | ✓ |  | ✓ |  | ✓ |  |  |
| #18 | Non-PS |  |  |  |  |  |  |  |
| #19 | PS | ✓ | ✓ | ✓ | ✓ | ✓ | ✓ | ✓ |
| #19 | Non-PS |  |  |  |  |  | ✓ |  |
| #20 | PS | ✓ | ✓ |  |  |  |  |  |
| #20 | Non-PS |  |  |  |  |  |  |  |
| #21 | PS | ✓ | ✓ | ✓ | ✓ | ✓ | ✓ |  |
| #21 | Non-PS |  |  |  |  |  |  |  |
| #22 | PS | ✓ | ✓ |  | ✓ |  |  |  |
| #22 | PS: Telephone PS | ✓ | ✓ | ✓ | ✓ |  |  |  |
| #22 | Non-PS |  |  |  | ✓ |  |  |  |
| #23 | PS | ✓ | ✓ |  | ✓ | ✓ | ✓ |  |
| #23 | Non-PS |  |  |  |  |  |  |  |
| #23 | Non-PS |  |  |  |  |  |  |  |
| #24 | PS | ✓ |  |  | ✓ | ✓ | ✓ |  |
| #24 | Non-PS |  |  |  |  |  |  |  |
| #25 | PS | ✓ | ✓ |  | ✓ | ✓ |  |  |
| #25 | Non-PS |  |  |  |  |  |  |  |
| #26 | PS | ✓ | ✓ | ✓ |  |  |  |  |
| #26 | Non-PS |  |  |  |  |  |  |  |
| #27 | PS | ✓ |  |  |  |  |  | ✓ |
| #27 | Non-PS |  |  |  |  |  |  |  |
| #28 | PS | ✓ | ✓ | ✓ | ✓ |  | ✓ | ✓ |
| #28 | Non-PS |  | ✓ |  | ✓ |  |  |  |
| #28 | Non-PS |  |  |  |  |  |  |  |
| #29 | PS | ✓ | ✓ | ✓ | ✓ | ✓ | ✓ |  |
| #29 | Non-PS |  |  |  |  |  |  |  |
| #30 | PS | ✓ | ✓ | ✓ | ✓ | ✓ | ✓ |  |
| #30 | Non-PS |  |  |  |  |  |  |  |
| #31 | PS | ✓ | ✓ | ✓ | ✓ |  |  |  |
| #31 | Non-PS: Partner without psychiatric history | ✓ | ✓ | ✓ | ✓ |  |  |  |
| #31 | Non-PS |  |  |  |  |  |  |  |
| #32 | PS | ✓ | ✓ | ✓ | ✓ | ✓ |  | ✓ |
| #32 | Non-PS |  |  |  |  |  |  |  |
| #33 | PS: Assertive Community Treatment with PSW | ✓ | ✓ |  |  |  |  |  |
| #33 | Non-PS: Assertive Community Treatment with non-PSW | ✓ | ✓ |  |  |  |  |  |
| #33 | Non-PS |  |  |  |  |  |  |  |
| #34 | PS: Telephone PS | ✓ |  | ✓ |  | ✓ |  | ✓ |
| #34 | Non-PS: Telephone without PS |  | ✓ |  |  | ✓ |  |  |
| #34 | Non-PS |  |  |  |  |  |  |  |
| #35 | PS | ✓ | ✓ | ✓ | ✓ | ✓ | ✓ |  |
| #35 | Non-PS | ✓ |  |  |  |  |  |  |
| #36 | PS |  |  |  | ✓ |  | ✓ |  |
| #36 | Non-PS |  |  |  |  |  |  |  |

PS=Peer support. Where no further description is provided, ‘PS’ refers to a peer support intervention arm, and ‘Non-PS’ refers to care as usual. Peer support components were labelled from B onwards, as care as usual was coded as A in the analysis.
